# The health and economic repercussions of declining MMR coverage in the United States

**DOI:** 10.64898/2026.02.19.26346619

**Authors:** Chad R. Wells, Abhishek Pandey, Yang Ye, Carolyn Bawden, Rebecca Giglio, Charlene Wong, Velda Wang, Chelsea Cipriano, Lamia Ayaz, Gergely Röst, Seyed M. Moghadas, Meagan C. Fitzpatrick, Burton H. Singer, Alison P. Galvani

**Affiliations:** Center for Infectious Disease Modeling and Analysis (CIDMA), Yale School of Public Health, New Haven, Connecticut, 06510, USA; The Common Health Coalition, New York, NY, 10016; National Laboratory for Health Security, University of Szeged, Szeged, Hungary; Agent-Based Modelling Laboratory, York University, Toronto, Ontario, M3J 1P3, Canada; Center for Vaccine Development and Global Health, University of Maryland School of Medicine, Baltimore, Maryland, 21201, USA; Emerging Pathogens Institute, University of Florida, Gainesville, Florida, 32610, USA

**Keywords:** childhood immunization, measles, outbreak, economic burden

## Abstract

The resurgence of measles in the United States, driven by declining childhood vaccination coverage, poses a substantial public health and economic threat. Using county-level MMR vaccine coverage data and spatial incidence models, we quantified the economic burden of measles in 2025 and projected the impact of continued declines in vaccine uptake. In 2025, the estimated cost per measles case was $104,629 (50% High-Density Interval [HDI]: $100,729–$110,140), yielding a national burden of $244.2 million (50% HDI: $69.9–$872.5 million). The cost per case varied widely across counties and was inversely correlated with local population immunity levels (Spearman correlation = -0.75, *p* < 0.001). We modeled a scenario in which coverage among children aged 0–6 years declined by 1% per year, reaching a 5% absolute reduction by year 5 relative to baseline. Under this scenario, we projected a nonlinear surge in cases, hospitalizations, and annual expenditures arising from outbreak response, direct medical costs, and productivity losses. This scenario produced 17,232 (50% HDI: 9,177–26,428), 4,085 (50% HDI: 2,184–6,210) hospitalizations, 36 (50% HDI: 19–54) deaths, and $1.50 (50% HDI: $0.90–$2.85) billion in annual costs in 2030, with a cumulative cost of $7.77 billion (50% HDI: $5.56–$11.58 billion) over 5 years. These findings demonstrate that even marginal reductions in MMR vaccine uptake can result in disproportionately large health and economic burdens.

**Significance Statement:** The United States is experiencing a resurgence of measles amid recent declines in childhood MMR vaccination. Using mathematical modeling informed by spatially resolved data on vaccination coverage, incidence, and associated economic costs, we quantified both the current and projected financial burden of measles in the United States under continued declines in coverage. For 2025, we estimated that measles imposes a cost of $244.2 million nationwide, with substantial heterogeneity in cost per case across counties driven by gaps in population immunity. Even modest annual reductions in vaccine coverage among young children generate a nonlinear increase in cases and hospitalizations, with costs totaling $7.77 billion over a five-year period.

## Introduction

Childhood immunization has been one of the most effective public health interventions for reducing morbidity and mortality from infectious diseases (1). However, recent policy shifts in the United States (US) threaten to undermine this achievement. Policies constraining healthcare funding, altering eligibility for public insurance programs, or disrupting vaccination outreach and delivery systems may collectively reduce routine childhood vaccination rates (2, 3). On January 5, 2026, the US Centers for Disease Control and Prevention announced changes to the vaccine schedule in the absence of new scientific evidence (4, 5), a step that may undermine confidence not only in the targeted vaccines but in vaccination more broadly. Similarly, cuts to Medicaid funding and healthcare subsidies are likely to restrict access to vaccines, because clinical visits often involve fees even when vaccines are provided at no cost, forcing parents who have lost health insurance to pay out of pocket or forgo care. Even modest reductions in vaccine coverage can have profound consequences for population-level immunity, particularly for highly transmissible diseases such as measles (6, 7).

Declines in Measles-Mumps-Rubella (MMR) vaccination coverage not only increase the probability of outbreaks but also amplify their size, duration, and geographic spread (8–10). The recent resurgence of measles in regions where it was previously considered eliminated underscores the fragility of protection against infectious disease (11). Over the last year in the US, outbreaks with epicenter in Texas, Utah, and South Carolina have posed a substantial risk to national elimination status (12).

As measles has been rare in the US for the past three decades, its symptoms may not be widely recognized, contributing to underreporting. While a mild case may include the characteristic measles rash of flat red spots accompanied by other nonspecific acute viral symptoms (13), approximately 20% of cases will progress to higher severity (14). These more severe manifestations can include pneumonia, febrile seizures, acute disseminated encephalomyelitis, subacute sclerosing panencephalitis, and death (15, 16). Hospitalization is a common consequence of severe measles. In 2025, 19% of reported measles cases among children younger than 5 years required hospitalization (17). Case fatality rates are disproportionately elevated among infants and older adults (18, 19). Beyond the immediate health burden measured in increased hospitalizations, severe cases, and deaths, measles outbreaks impose substantial economic costs on health systems and society (20, 21). These include direct medical expenditures, costs of public health response, and productivity losses. However, the full magnitude of these impacts has not been systematically quantified at the national level in the US. Moreover, societal costs associated with measles outbreaks, including productivity losses from missed school and work, have not been comprehensively assessed (22, 23).

To evaluate the repercussions of potential declines in MMR coverage, we developed an integrated modeling framework that propagates county-level changes in childhood vaccination coverage to outbreak risks and economic consequences across the US. Our analyses provide a comprehensive assessment of the health and economic repercussions of US policies that may lead to a reduction in vaccination uptake.

## Results

We first inferred county-level MMR vaccine coverage using our regression framework, which integrates reported kindergarten coverage, socio-demographic covariates, and insurance type (**Methods**). The model was trained on county- and state-level MMR coverage from the 2017–2018 through 2023–2024 school years, with separate logits for privately insured, publicly insured, and uninsured children. For counties without kindergarten coverage data, we extrapolated MMR coverage using the fitted regression coefficients, applying state-specific adjustment factors to ensure agreement with reported state-level coverage (**Figure S1**). These estimates reveal clusters of counties with lower coverage among children aged 0–6 years in the East North Central and Mid-Atlantic regions, as well as sporadic pockets across the Western US (**Figure S1F**).

To quantify measles burden, we combined county-level vaccination coverage with age-specific contact matrices and a measles transmission model. Importation events were modeled using international measles importation patterns observed in 2025 (24), and onward spatial spread was simulated through a gravity framework capturing inter-county connectivity. County-level outbreak sizes were then aggregated to generate national estimates (**Figure S2A)**. We applied costs associated with outbreak response, direct medical costs, and productivity losses to estimate the overall economic burden (**Table S1**).

Under the baseline MMR coverage, we estimated 2,181 measles cases (50% High-Density Interval [HDI]: 719–8,176), 554 hospitalizations (50% HDI: 196–1,927), and 5 deaths (50% HDI: 2–17), corresponding to a national economic burden of $244.2 million (50% HDI: $69.9–$872.5 million) with a cost per case of $104,629 (50% HDI: $100,729–$110,140) in 2025 (**Table S2)**. Within individual ensembles, outbreak response activities, including contact tracing, testing, and post-exposure vaccination, accounted for the largest share of total costs (65.21%; 50% HDI: 62.50%–66.92%), followed by productivity losses due to missed work and school (32.07%; 50% HDI: 30.11%–34.80%), then direct medical expenditures (2.95%; 50% HDI: 2.66%–3.16%) (**Table S3)**. Across all samples, the cost of outbreak response activities was $144.3 million (50% HDI: $44.2–$556.5 million), productivity losses were $84.4 million ($23.0–$286.2 million), and $5.9 million (50% HDI: $2.5–$24.4 million) for the direct medical costs. Within the direct medical expenditures associated with outpatient visits and hospitalizations, $3.01 million (50% HDI: $1.43–$14.92 million) was borne by those with private insurance, $0.68 million (50% HDI: $0.35–$3.27 million) by those with public insurance, and $1.87 million (50% HDI: $0.60–$5.69 million) by uninsured individuals (**Table S2–S3**).

### County-level variation in cost per case

The economic impact of measles outbreaks closely reflected county-level variation in MMR coverage (**Figure S1**). We calculated the cost per case for each county, assuming a fixed outbreak size within a county. Across counties, the median cost per case was $85,637, with a standard deviation of $12,418 and an interquartile range of $80,130–$92,352 (**Figure 1**). Cost per case was strongly and inversely correlated with population-level immunity against measles (Spearman correlation: -0.75; *p* < 0.001), indicating that counties with the lowest immunity not only faced higher outbreak risks but also incurred substantially greater cost per case. Higher local wages and hospitalization costs also contributed to elevated cost per case.

**Figure 1.**
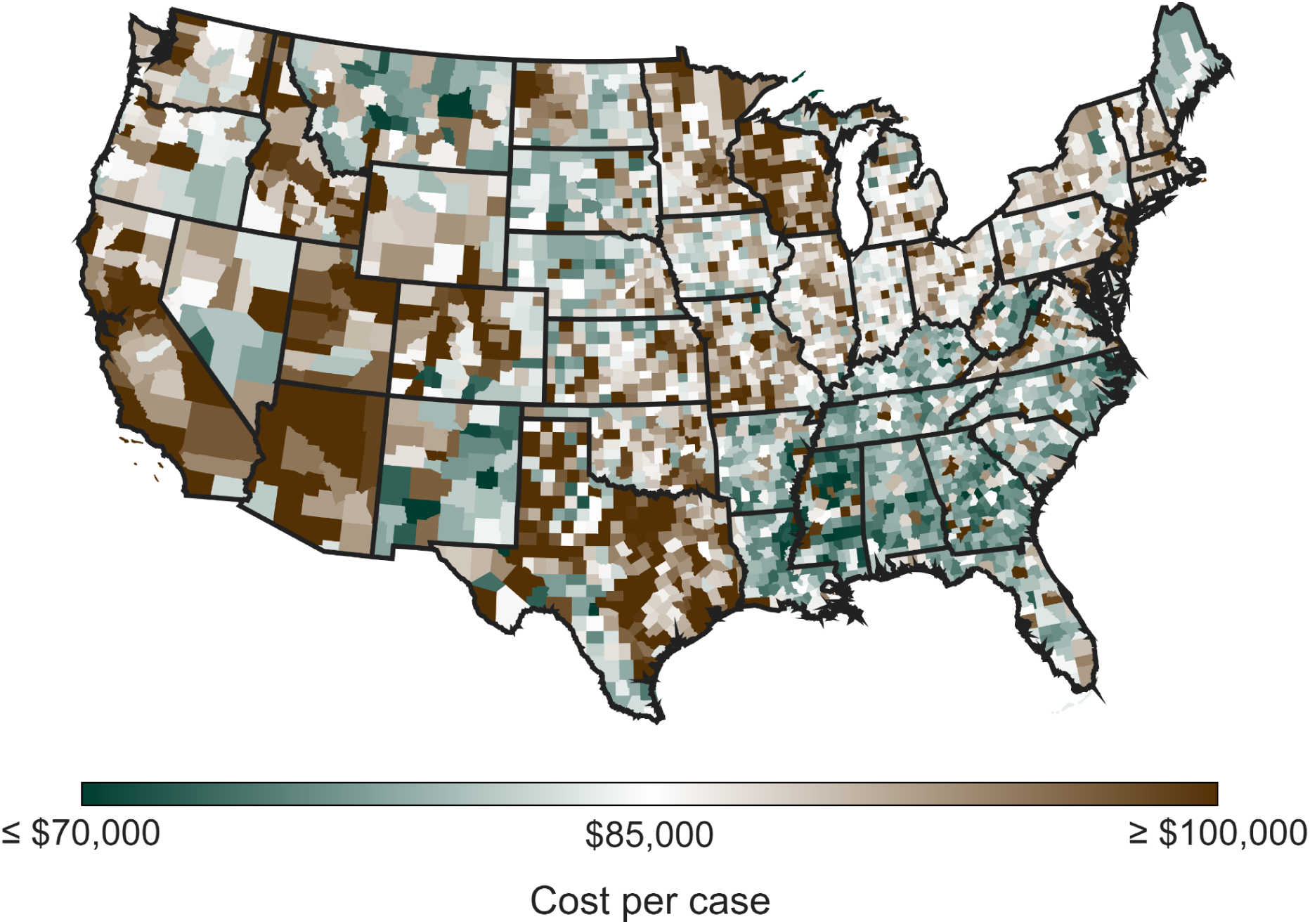
County-level estimates of cost per measles case. Mode of the estimated cost per case by US county, assuming a fixed outbreak size of 50 cases in the county.

### Health and economic impacts of declining MMR coverage

To assess the impact of continued declines in childhood MMR coverage on measles burden, we simulated scenarios in which national coverage among children aged 0–6 years decreased by 0.25%, 0.5%, 0.75%, or 1% per year over five years, beginning in 2026. Under each scenario, county-level immunity profiles were updated annually using an age-structured exponential decline in vaccination coverage among affected cohorts, while immunity in older age groups (≥15 years) and measles importation patterns were held constant at baseline levels. Using the same importation structure as in 2025, we then projected annual measles incidence and associated costs from 2026 to 2030.

Under the scenario of a 1% absolute annual reduction in MMR coverage among children aged 0–6 years relative to 2025 levels (**Figure S1**), measles burden was projected to increase over time (**Table 1 and Table S4-S8),** reaching 17,232 (50% HDI: 9,177–26,428) cases, 4,085 (50% HDI: 2,184–6,210) hospitalizations, and 36 (50% HDI: 19–54) deaths in 2030. This represents an over sevenfold increase in cases, hospitalizations, and deaths relative to the estimated baseline in 2025. Corresponding annual costs would increase to $1.50 billion (50% HDI: $0.90–$2.85 billion) in 2030, an over sixfold rise from costs in 2025 (**Table 1**). Over the five-year period, cumulative national costs attributable to measles outbreaks would reach $7.77 billion (50% HDI: $5.56–$11.58 billion) (**Table S8**).

**Table 1.** Annual measles-attributable costs (in millions USD) in the US under declining MMR vaccination coverage. Mode (and 50% HDI) annual costs associated with measles cases, assuming 2025 importation patterns, under absolute annual reductions from 0.25% to 1% in MMR vaccination coverage among children aged 0–6 years.

| Year | Annual reduction in MMR vaccination coverage among children aged 0–6 years |  |  |  |
| --- | --- | --- | --- | --- |
|  | 0.25% | 0.5% | 0.75% | 1% |
| 2025 | \$244.2<br>(\$69.9–\$872.5) | \$244.2<br>(\$69.9–\$872.5) | \$244.2<br>(\$69.9–\$872.5) | \$244.2<br>(\$69.9–\$872.5) |
| 2026 | \$253.3<br>(\$73.9–\$893.5) | \$258.4<br>(\$76.2–\$912.6) | \$268.8<br>(\$80.7–\$934.9) | \$282.1<br>(\$85.6–\$959.5) |
| 2027 | \$260.8<br>(\$77.3–\$914.7) | \$291.7<br>(\$86.4–\$966.0) | \$324.5<br>(\$98.1–\$1,028.4) | \$440.2<br>(\$111.5–\$1,102.6) |
| 2028 | \$280.5<br>(\$84.6–\$949.1) | \$349.9<br>(\$106.0–\$1,065.8) | \$540.8<br>(\$140.2–\$1,227.8) | \$613.9<br>(\$234.4–\$1,452.5) |
| 2029 | \$311.9<br>(\$95.4–\$1,007.9) | \$541.6<br>(\$150.2–\$1,248.6) | \$667.4<br>(\$298.5–\$1,592.6) | \$1,009.2<br>(\$512.6–\$2,017.8) |
| 2030 | \$371.8<br>(\$111.7–\$1,093.9) | \$630.2<br>(\$279.9–\$1,544.4) | \$1,088.4<br>(\$551.0–\$2,122.0) | \$1,498.5<br>(\$901.2–\$2,852.4) |

Smaller annual reductions in MMR coverage also generated progressively smaller, but still substantial, increases in health and economic burdens (**Table 1 and Table S8**). Even under a moderate annual coverage decline of 0.5%, hospitalizations were projected to rise to 1,432 (50% HDI: 726–3,420) and deaths to 12 (50% HDI: 6–30) in 2030. In parallel, reductions in coverage increased the cost per measles case, largely due to rising costs for outbreak response and productivity losses. By 2030, a 1% annual reduction in MMR coverage was projected to increase the cost per case by $3,094 relative to 2025 (**Table S7**).

### Impact of importation events on national incidence

The projected number of national measles cases increased with the number of importation events. In observed data, 170 importations in 2025 were associated with 2,181 (50% HDI: 719–8,176) total measles cases, compared with 53 importations and 80 (50% HDI: 56–368) total cases in 2024, and 13 importations and 15 (50% HDI: 13–53) total cases in 2023 (**Table S2**).

To further quantify this relationship, we conducted a scenario analysis in which the annual number of measles importations ranged from 25 to 200 events per year. Importations were seeded randomly across counties in proportion to population size. We found that annual measles incidence (i.e., including both domestic and imported cases) increased monotonically with the number of importation events (**Figure 2A**), with a total of 33 (50% HDI: 26–79) cases arising from 25 importations and 513 (50% HDI: 319–11,357) cases from 200 importations. As importations increased, uncertainty in total case counts also increased, reflecting the higher probability that one or more importations would trigger a large outbreak.

**Figure 2.**
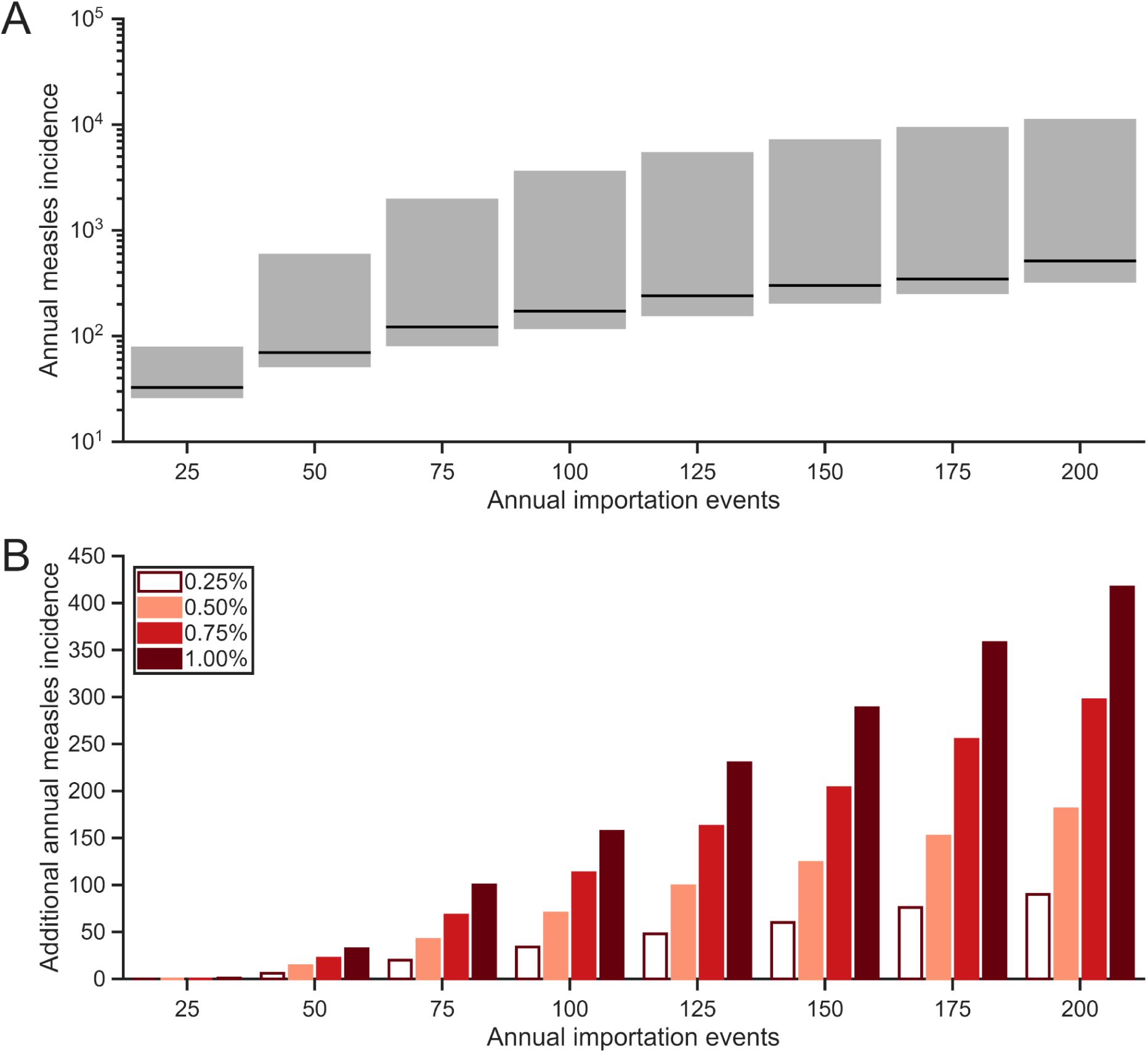
Influence of measles importations on annual incidence. Assuming imported measles cases are randomly distributed into counties proportional to their population size, panels show: (A) mode (solid line) and 50% high-density interval (gray area) of annual measles cases (i.e., domestic and imported) under baseline vaccination coverage (in 2025) as a function of the number of importation events; and (B) median additional annual measles cases associated with absolute reductions of 0.25% (white), 0.50% (light red), 0.75% (red), and 1% (dark red) in MMR vaccination coverage among children aged 0–6 years.

The interaction between importation pressure and reduced vaccination coverage further amplified measles burden (**Figure 2B**). For example, under 25 annual importation events, a 0.25% absolute reduction in MMR coverage among children aged 0–6 years resulted in a median of zero additional annual cases, increasing to a median of one additional case for a 1% absolute reduction. In contrast, 200 annual importation events generated a median of 90 additional cases for a 0.25% absolute reduction and 417 additional cases for a 1% absolute reduction in MMR coverage. These results indicate that in the context of rising global measles incidence, maintaining high vaccination coverage is critical to limit domestic measles burden in the US.

## Discussion

MMR vaccination coverage among US kindergarteners has declined steadily since the 2019–2020 school year, coinciding with the resurgence of measles nationally. In 2025, the US experienced its largest number of annual measles cases since 1992 and approached the threshold for losing measles elimination status (25, 26). We estimated that measles outbreaks in 2025 incurred $244.2 million in societal costs, corresponding to $104,629 per case. These costs were highly heterogeneous across counties and strongly inversely associated with the level of population immunity. Our results further indicate that even modest annual reductions in MMR coverage among children aged 0–6 years could lead to sharp increases in hospitalizations and costs, with a sustained 1% annual decline in coverage generating an additional $7.77 billion in measles-related costs over five years.

The economic burden of measles in 2025 was dominated by outbreak response activities ($144.3 million), followed by productivity losses ($84.4 million) and direct medical costs ($5.9 million). Although these estimates exceed those reported in prior studies with similar case counts (27), this difference reflects our broader societal perspective, which explicitly incorporates productivity losses, including school absenteeism. Most prior cost analyses have focused primarily on public health and medical expenditures, with limited attention to the broader societal impacts of measles outbreaks. Accounting for these wider costs provides a more complete assessment of the consequences of declining MMR coverage at both regional and national scales.

Our results nonetheless likely underestimate the true economic and societal toll of measles outbreaks. While we incorporated productivity losses from school absenteeism, we did not include losses in the human capital associated with missed learning (28–30). For every 10 days of absence per school year, lifetime earnings may decrease by up to 2% (28). In addition, our analysis focuses on costs arising from acute infection, as do most others, excluding rare long-term sequelae. Subacute sclerosing panencephalitis (SSPE), for example, is a progressive complication that manifests years after infection, with the highest risk among infants (1 per 609 cases) and children under five (1 per 1367 cases) (16, 31). Acute disseminated encephalomyelitis occurs in 1 per 1000 cases among older children and within two to four weeks of infection (15, 16). Across the subtypes of measles-associated encephalitis, the estimated lifetime societal cost exceeds $2.5 million (2025 USD) per case (32). Accounting for this outcome would increase the expected cost per unvaccinated measles case by more than $2,500. Those at the other end of the age distribution are also disproportionately impacted since the elderly are more likely to be immunocompromised, which in turn elevates risk of severe measles complications. Immunocompromised individuals are particularly at risk of fatal outcomes such as giant-cell pneumonia and measles inclusion body encephalitis, which may manifest months after infection. Declining MMR coverage among children increases outbreak risk while the overall population is aging and becoming increasingly vulnerable.

We also excluded the societal cost of measles-related deaths by not applying a value of statistical life lost, given ongoing debate regarding its interpretation across different age groups (33). If the Department of Health and Human Services value of $13,913,479 (2025 USD) per life lost were applied (34), estimated costs would increase by $69.6 million for 2025, and by as much as $500.9 million for 2030. These omissions indicate that our estimates are conservative and further underscore the long-term economic value of MMR vaccination.

We identified pronounced county-level heterogeneity in the cost per measles case, with higher costs in areas having lower population immunity. Although similar heterogeneity has been previously reported (22, 35), our analysis provides a mechanistic explanation: declining immunity increases the likelihood of severe disease, expands the scope of contact tracing, quarantine and isolation, and necessitates additional emergency vaccination of susceptible contacts. These dynamics inflate per-case costs even across counties with otherwise similar outbreak probability, highlighting that maintaining high MMR coverage reduces not only measles incidence but also the economic severity of outbreaks.

For this reason, the reporting of cost per measles case as a normalized metric can obscure any substantial heterogeneity existing in underlying epidemiological and demographic conditions. Our county-level analysis demonstrates that variation in immunity levels, contact patterns, and population structure alone can produce large differences in per-case costs, particularly for direct medical expenditures in small outbreaks where hospitalizations may be infrequent. Interpretation of cost-per-case estimates therefore requires careful consideration of the local context in which outbreaks occur.

Recent trends suggest that MMR coverage may continue to decline. Since the 2022–2023 school year, kindergarten MMR coverage has dropped by approximately 0.3–0.4 percentage points annually (36). Concomitantly, the US is experiencing a decline in MMR coverage among children aged 13 months (37), the age for which a first dose of MMR vaccination is recommended. Policy changes, proposed legislation, and shifts in the social environment may further erode coverage in coming years (38–41). Although our five-year projections in the reduced vaccination coverage scenario may overestimate long-term burden by not accounting for the accumulation of natural immunity, longer-term analyses indicate that a 10% relative decline in MMR coverage among young children could result in 11.1 million measles cases over 25 years (6). Applying our 2025 cost estimates implies average annual costs exceeding $46 billion under such a scenario. Even over shorter horizons, our results demonstrate that annual coverage declines of just 0.25%–1% produce rapidly compounding health and economic burdens. While recent changes to immunization recommendations have not affected MMR directly (4, 42), increased barriers to healthcare access may indirectly suppress uptake, increasing vulnerability to future outbreaks of measles and other vaccine-preventable diseases.

Our importation analyses further capture how declining domestic immunity amplifies the impact of measles introductions from abroad. Global measles incidence has increased in recent years because of suboptimal vaccine coverage worldwide (43, 44). Although international initiatives such as the Global Alliance for Vaccines and Immunization (GAVI) have improved access to vaccines for children in low-income countries and substantially reduced measles incidence worldwide (45, 46), funding shortfalls threaten these gains, including a $3 billion gap in 2025 partly due to reduced US contributions (47). Many US measles importations originate from regions where GAVI-supported programs have been active, including Eastern Mediterranean, Africa, and South East Asia (48, 49). The recent loss of measles elimination status in Canada further heightens importation risk (11). In contrast, countries with sustained high MMR vaccination coverage, such as Hungary, have avoided large outbreaks despite regional transmission. For example, in 2024, Hungary experienced only 18 despite high mobility and significant daily commuting between Romania and Austria, which had 30,692 and 542 cases respectively (50)

Our model estimated higher hospitalizations than those reported for 2025, which may reflect variability in reporting practices compared with previous years from which hospitalization rates were derived. For instance, hospitalization estimates for 2024 included admissions for measles isolation as well as clinical complications, yielding a reported hospitalization rate of approximately 40%, whereas 2025 reports appear to reflect hospitalizations primarily for disease complications, with a lower reported rate of 11% (17). When age- and vaccination-specific rates for severe disease were applied (14), hospitalization estimates aligned more closely with the observed 2025 data. However, uncertainty in hospitalization rates had limited influence on overall costs, which were driven primarily by outbreak response activities and productivity losses.

Our study has several limitations. Importation events were simulated based on observed patterns and generated randomly and proportional to population size; however, this approach may not fully capture the complexity of international mobility and the dynamic, interconnected nature of global measles transmission. In the absence of region-specific data on the cost of the public health response, we assumed a homogeneous cost across counties. While this assumption may affect absolute cost estimates, it preserves the relative comparison of counties within a state and identifies those likely to incur the largest cost burden. Finally, we did not account for broader social disruptions or healthcare system constraints, which could further amplify the impact of large outbreaks.

In summary, the resurgence of measles in 2025 reflects the combined effects of declining vaccination coverage and increased importation pressure. Between 1994 and 2023, measles vaccination prevented an estimated 104 million cases and 85,000 deaths in the US (51). Continued erosion of MMR coverage threatens to reverse these gains, imposing substantial and rapidly escalating health and economic costs. Sustained investment in vaccination policy, public health infrastructure, and global immunization efforts is therefore paramount to protect population health and alleviate the socioeconomic impact of measles.

## Methods

### Estimation of county-level MMR vaccine coverage

We estimated county-level MMR vaccination coverage among kindergarten-aged children using an insurance-stratified logistic regression framework. The model incorporated demographic, socioeconomic, and policy-related covariates, including exemption policies, population-to-physician ratios, household income, and educational attainment. Separate logits were specified for privately insured, publicly insured, and uninsured children to capture heterogeneity in access and uptake.

The model was fit and validated using reported county-, state-, and Nebraska health district-level school-entry coverage data from the 2017–2018 through 2023–2024 school years under a likelihood approach. To ensure consistency with observed coverage, model intercepts were calibrated so that the aggregated county-level predictions matched reported county- or state-level MMR coverage for the 2023–2024 school year. To estimate MMR coverage in additional age groups (0–4, 10–14, 15–19, and 20–24 years), we derived state-level adjustment factors using the Centers for Disease Control and Prevention (CDC) survey data and applied these uniformly across counties within each state (**SI Appendix**).

### Construction of county-level immunity profiles

We translated age-specific vaccination coverage into immunity profiles using the routine two-dose MMR schedule. Children under five years of age were assumed to have received only the first dose, with 93% vaccine efficacy protection against infection (52), while the second dose was assumed to be administered at age five, providing an improved protection of 97% (52). For individuals aged 25 years or older, we applied previously estimated state-level measles immunity uniformly across counties within each state (6). Combining age-specific coverage, vaccine efficacy, and natural immunity, we generated an age-structured immunity profile for each county, which served as inputs for transmission modeling.

### Modeling measles importation and spatial spread

Annual measles incidence was driven by both international importation events and domestic spatial spread between counties. The probability that an imported case initiated local transmission followed a negative binomial process. Between-county spread was modeled using a gravity structure, with connectivity proportional to county population sizes and inversely related to geographic distance. Together, these processes defined a hurdle structure (53) consisting of (i) the probability that a county experienced zero cases and (ii) a positive component describing the size distribution of outbreaks conditional on transmission occurring.

### Within-county transmission dynamics

Measles transmission within a county was modeled using a combination of an age-structured final-size framework and a hurdle model (**SI Appendix**) (54). For each county *c*, we computed the effective reproduction number *R_E,c_*, which depended on the county-specific immunity profile, state-level age-specific contact matrices (55), and a county-specific effective transmission rate.

The county-specific effective transmission rate was assumed to depend on county-level demographic characteristics, including median family income, the GINI index, population stratified by education, population stratified by poverty-income ratio, and rural-urban continuum code (**SI Appendix**).

When *R_E,c_* > 1, we solved the standard age-structured final-size equations to estimate age-specific attack rates and total outbreak size (54). When *R_E,c_* ≤ 1, transmission was approximated using a stuttering-chain branching process (56), with the expected chain size capped at 100 cases (7). We trained the model to national and geographic patterns of cumulative measles incidence reported as of December 31, 2025 (**SI Appendix**).

### Estimation of economic costs

Economic impacts were estimated by linking simulated cases and contacts to three cost components: (i) public health response costs (i.e., contact tracing, testing, and post-exposure vaccination); (ii) direct medical costs (i.e., outpatient care and hospitalization); and (iii) productivity losses due to missed work and school. Cost inputs were drawn from national and state-specific data sources and expressed in 2025 US dollars. Where applicable, costs were further stratified by payer type (i.e., private insurance, public insurance, uninsured) (**SI Appendix and Table S1**).

### Scenarios of declining MMR vaccination coverage

To evaluate the impact of continued declines in childhood MMR coverage, we simulated annual age-dependent reductions confined to children aged 0–6 years, with the effect propagating to older age groups over a five-year period. Reductions in coverage for the 0–4, 5–9, and 10–14 year-olds were approximated using an age-dependent exponential decline from baseline vaccine coverage (**SI Appendix**). For each scenario, county-level coverage estimates were recalibrated by adjusting the logistic transform to achieve the specified national reduction (**SI Appendix**). County-specific effective reproduction numbers under the reduced vaccine coverage were recomputed annually, and measles incidence was simulated under the updated vaccination profiles.

### Importation assumptions and scenario analyses

For scenarios involving reduced vaccine coverage, we assumed that the importation events were equivalent to those reported in 2025. For 2023 and 2024, when only state-level importation locations were available, imported cases were randomly assigned to counties within each state proportional to county population size. In additional scenario analyses, a fixed number of annual importation events was randomly seeded into counties nationwide based on population size to examine the sensitivity of national incidence to importation intensity.

### Uncertainty and reproducibility

All results are reported as the mode and 50% high-density interval (HDI) from 2,500 Monte-Carlo samples (**SI Appendix**). The 50% HDI is defined as the narrowest interval containing 50% of the simulated probability mass around the mode. Computational code used to run simulations and conduct analyses is available in an online repository (57).

## Data Availability

All data produced in the present work are contained in the manuscript and available online at https://github.com/WellsRC/Measles_USA.

## Funding acknowledgment

SMM Acknowledges the support from the Natural Sciences and Engineering Council of Canada Discovery and Alliance Grants, and the Ontario Research Fund, Research Excellence. This study was in part supported by HU-RIZONT international research excellence programme (Rapid-GRIP project: 2024-1.2.3-HU-RIZONT-2024-00034).

