## Supporting Information for "The health and economic repercussions of declining MMR coverage in the United States"

**Supplementary Methods** **2**
County-level vaccine coverage 2
Simulated annual measles cases 4
Parameterization of vaccine coverage model 8
Parameterization of annual measles cases 10
Reduction in MMR vaccine coverage 13
Hospitalization, Severe Disease, and Death 14
Costs 14
Monte Carlo Samples 15
**Supplementary References** **16**
**Supplementary Figures** **19**
**Supplementary Tables** **21**

### Supplementary Methods

*County-level vaccine coverage*

For the vaccine coverage from five to nine years of age, we utilized county-level MMR vaccine coverage among kindergarteners. To provide county-level estimates of vaccine coverage among kindergarteners for counties that did not report data, we trained a series of logistic regression models using available county-level vaccination coverage data from the 2017–18 school year through the 2023–24 school year (**File S1**). The covariates explored were the allowance of religious exemptions, the allowance of philosophical exemptions, the number of people per doctor (1), age (2), race (2), median household income (2), Gini index (2), level of education (2), poverty income ratio (2), and rural-urban code (3).

We identified the best predictive model based on its spatial validation capabilities and ability to fit the training data. In each model, we included the covariates of vaccine exemptions and population per doctor to explore the effects of access to care and policy changes in vaccine exemptions. We used the US Census region division to stratify counties for the spatial validation.

For each combination of covariates explored, there consisted of three logistic regression models which provided an estimate of the vaccine coverage among kindergarteners who were uninsured in county $c$

$$\log\left(\nu_{U,c}/\left(1-\nu_{U,c}\right)\right)=\beta_U+\sum_{j=1}^{N}\beta_j X_{j,c}+\sum_{i=1}^{N}\sum_{j=i}^{N}\beta_{i,j}X_{i,c}X_{j,c}, \quad [1]$$

the vaccine coverage among kindergarteners who are privately insured in county $c$

$$\log\left(\nu_{P,c}/\left(1-\nu_{P,c}\right)\right)=\beta_P+\sum_{j=1}^{N}\beta_j X_{j,c}+\sum_{i=1}^{N}\sum_{j=i}^{N}\beta_{i,j}X_{i,c}X_{j,c}, \quad [2]$$

and the vaccine coverage among kindergarteners who had publicly insured in county $c$

$$\log\left(\nu_{M,c}/\left(1-\nu_{M,c}\right)\right)=\beta_M+\sum_{j=1}^{N}\beta_j X_{j,c}+\sum_{i=1}^{N}\sum_{j=i}^{N}\beta_{i,j}X_{i,c}X_{j,c}. \quad [3]$$

The overall vaccine coverage among kindergarteners in a county is calculated as

$$\nu_c=q_{U,c}\nu_{U,c}+q_{P,c}\nu_{P,c}+q_{M,c}\nu_{M,c}, \quad [4]$$

where $q_{U,c}$ is the proportion of the population under the age of 6 years who are uninsured in county $c$, $q_{P,c}$ is the proportion of the population under the age of 6 years who are privately insured in county $c$, and $q_{M,c}$ is the proportion of the population under the age of 6 years who are publicly insured in county $c$. We do not consider the interaction terms that include the rural-urban code, allowance of religious exemptions, or allowance of philosophical exemptions in the construction of the model. We note that the proportion of the population who are privately insured is data informed by Census data measuring those with private insurance or in combination. Similarly, the proportion of the population who are publicly insured is data informed by Census data measuring those with public insurance or in combination. As a result there is some overlap between, resulting in some cases where $q_{U,c} + q_{P,c} + q_{M,c} \neq 1$. To mitigate this issue, we normalized the values such that they summed to one.

For counties in which the vaccine coverage among kindergarteners is reported, we adjust the logit transformed values from the trained model by $\Delta z_c$

$$\log\left(\hat{\nu}_{U,c}/\left(1 - \hat{\nu}_{U,c}\right)\right) = \beta_U + \sum_{j=1}^{N} \beta_j X_{j,c} + \sum_{i=1}^{N}\sum_{j=i}^{N} \beta_{i,j} X_{i,c} X_{j,c} + \Delta z_c \quad [5]$$

$$\log\left(\hat{\nu}_{P,c}/\left(1 - \hat{\nu}_{P,c}\right)\right) = \beta_P + \sum_{j=1}^{N} \beta_j X_{j,c} + \sum_{i=1}^{N}\sum_{j=i}^{N} \beta_{i,j} X_{i,c} X_{j,c} + \Delta z_c \quad [6]$$

$$\log\left(\hat{\nu}_{M,c}/\left(1 - \hat{\nu}_{M,c}\right)\right) = \beta_M + \sum_{j=1}^{N} \beta_j X_{j,c} + \sum_{i=1}^{N}\sum_{j=i}^{N} \beta_{i,j} X_{i,c} X_{j,c} + \Delta z_c \quad [7]$$

such that

$$q_{U,c}\hat{\nu}_{U,c} + q_{P,c}\hat{\nu}_{P,c} + q_{M,c}\hat{\nu}_{M,c} = \hat{\nu}_c \quad [8]$$

where $\hat{\nu}_c$ is the reported vaccine coverage among kindergarteners in county $c$ for the 2023–2024 school year.

For each county in state $s$ where we do not have county-level vaccine coverage, we used a state-wide adjustment factor $\Delta y_s$ for the county-level logit transformed values from the trained model

$$\log\left(\hat{\nu}_{U,c}/\left(1 - \hat{\nu}_{U,c}\right)\right) = \beta_U + \sum_{j=1}^{N} \beta_j X_{j,c} + \sum_{i=1}^{N}\sum_{j=i}^{N} \beta_{i,j} X_{i,c} X_{j,c} + \Delta y_s \quad [9]$$

$$\log\left(\hat{\nu}_{P,c}/\left(1 - \hat{\nu}_{P,c}\right)\right) = \beta_P + \sum_{j=1}^{N} \beta_j X_{j,c} + \sum_{i=1}^{N}\sum_{j=i}^{N} \beta_{i,j} X_{i,c} X_{j,c} + \Delta y_s \quad [10]$$

$$\log\left(\hat{\nu}_{M,c}/\left(1 - \hat{\nu}_{M,c}\right)\right) = \beta_M + \sum_{j=1}^{N} \beta_j X_{j,c} + \sum_{i=1}^{N}\sum_{j=i}^{N} \beta_{i,j} X_{i,c} X_{j,c} + \Delta y_s \quad [11]$$

such that the state-level coverage is equivalent to that reported for the state. For states that did not report the vaccine coverage, we used the inferred value from the trained model, i.e. $\Delta y_s = 0$.

To estimate the county-level vaccine coverage among those under five years of age, 10 years to 14 years of age, 15 years to 19 years of age, and 20 years to 24 years of age, we estimated a an age-specific state-level adjustment factor $\Delta\tilde{y}_{s,a}$ for the regression model used to infer the vaccine coverage among kindergarteners

$$\log\left(\tilde{\nu}_{U,c}/\left(1-\tilde{\nu}_{U,c}\right)\right)=\beta_U+\sum_{j=1}^{N}\beta_jX_{j,c}+\sum_{i=1}^{N}\sum_{j=i}^{N}\beta_{i,j}X_{i,c}X_{j,c}+\Delta\tilde{y}_{s,a} \quad [12]$$

$$\log\left(\tilde{\nu}_{P,c}/\left(1-\tilde{\nu}_{P,c}\right)\right)=\beta_P+\sum_{j=1}^{N}\beta_jX_{j,c}+\sum_{i=1}^{N}\sum_{j=i}^{N}\beta_{i,j}X_{i,c}X_{j,c}+\Delta\tilde{y}_{s,a} \quad [13]$$

$$\log\left(\tilde{\nu}_{M,c}/\left(1-\tilde{\nu}_{M,c}\right)\right)=\beta_M+\sum_{j=1}^{N}\beta_jX_{j,c}+\sum_{i=1}^{N}\sum_{j=i}^{N}\beta_{i,j}X_{i,c}X_{j,c}+\Delta\tilde{y}_{s,a} \quad [14]$$

such that the estimated state-level coverage is equivalent to that reported for the state. We applied age-specific insurance rates for each county when computing overall county-level coverage for these other age groups. Specifically, we used the rates reported for those under the age of six years for the under five years of age, the rates reported for those six years to 18 years of age for the groups 10 years to 14 years of age and 15 to 19 years of age, and the rates reported for those 19 to 25 years of age for the group 20 years to 24 years of age.

The state-level MMR vaccine coverage among the children under the age of five years was computed using a weighted average of the vaccine coverage at 13 months (2021 birth year), 24 months (2021 birth year), 35 months (2021 birth year), and 35 months (2020 birth year).

$$\nu_{0-4}=\left(\nu_{13m}\omega_1+\nu_{24m}\omega_2+\omega_3\nu_{35m}+\omega_4\tilde{\nu}_{35m}\right)/\left(\omega_0+\omega_1+\omega_2+\omega_3+\omega_4\right) \quad [15]$$

where the age specified weights were determined using state-specific single age groups (4).

We used state-level vaccine coverage among those 13 to 17 years of age reported for 2023, 2021, and 2016 to approximate vaccine coverage among those 10 to 14 years of age, 15 to 19 years of age, and 20 to 24 years of age, respectively.

Given the absence of vaccination data among those 25 years of age or older, we needed to estimate the proportions of the population susceptible and vaccinated, as well as susceptible and unvaccinated, due to differences in hospitalization rates. For age groups 25 years of age and older, we used the model inferred values for the vaccine coverage. If the level of immunity exceeded that previous estimate (5), we implemented a correcting factor to reduce the vaccine coverage. If the level of immunity was below that of the previous estimate (5), the residual immunity was distributed as natural immunity.

*Simulated annual measles cases*

According to the child immunization schedule, children under four years of age have received only a single dose of a measles-containing vaccine, with an efficacy of 93% (6). The child immunization schedule recommends the second dose between the ages of four and six years. The efficacy after the second dose is 97% (6). To simplify estimating county-level vaccine-acquired immunity among children under five years of age, we assumed the second dose was administered at age five. For individuals 25 years and older, we used previous state-level model estimates of measles acquired immunity to each county (5).

For a specified transmission rate $\beta_c$, state-specific age contact matrix $M$ (7), immunity-profile $\vec{\varphi}_c$ in county $c$ and population size $\vec{N}_c$, we computed the effective reproduction number $R_{E,c}$ for each county. When the effective reproduction number $R_{E,c} > 1$ in county $c$ the final number of potential cases in county is

$$F_c = \sum_a N_{c,a}\left(1 - \varphi_{c,a}\right)z_a \qquad [16]$$

where $\vec{z}$ satisfies

$$A\vec{z} + \log\left(1 - \vec{z}\right) = \vec{0} \qquad [17]$$

where $\vec{0}$ is the zero-column vector and

$$A_{i,j} = \beta_c M_{i,j} \frac{N_{c,j}\left(1-\varphi_{c,j}\right)}{N_{c,j}} \qquad [18]$$

is the element of matrix $A$ in the $i^{th}$ row and $j^{th}$ column (8). We assumed that the county-level transmission rate was of the form

$$\log\left(\beta_c\right) = \sum_{j=1} \Upsilon_j X_{j,c} + \sum_{i=1}^{9} \Upsilon_{R,i} \mathcal{R}_{i,c}, \qquad [19]$$

where $\Upsilon_j$ is the coefficient for covariate $j$ and the covariates applied were the median family income, the GINI index, the proportion of the population with a specific education level (grade 9–12, high school graduate, some college, associate degree, bachelor degree, and graduate/professional degree), the proportion of the population with a specified poverty index ratio (under 0.5, 0.50–0.74, 0.75–0.99, 1.00–1.24, 1.25–1.49, 1.50–1.74, 1.75–1.84, 1.85–1.99, 2.00–2.99, 3.00–3.99, 4.00–4.99, 5.00 and over) and indicator variables for the rural urban continuum code (i.e. if the rural continuum code for county $c$ is $i$ then $\mathcal{R}_{i,c} = 1$; otherwise $\mathcal{R}_{i,c} = 0$ ). During initial model exploration, we found a relationship between the level of immunity in a county and the required transmission rate. Thus, we utilized selected components of the vaccination model to inform the county-level transmission rate. As age is integrated into the contact patterns, we did not include it as a covariate. We assumed that physicians per capita and race would not have a substantial influence on transmission or public health response (which would affect the effective reproduction number), thus, they were not included as covariates. In addition, we did not include the proportion of the population with less than grade 9 education because during initial exploration the covariance matrix of the coefficients was found to be singular (implying a covariate could be expressed as a linear combination of others).

When the effective reproduction number $R_{E,c} \leq 1$ in county $c$ the final number of potential cases in county is

$$F_c = \min\left\{\frac{1}{1-R_{E,c}}, 100\right\}, \qquad [20]$$

which is the average stuttering chain size (9). We assumed that a stuttering chain cannot exceed 100 cases (10).

We modeled the county-level case counts using a hurdle model, where the probability of observing zero annual measles cases in county $c$ is dependent on international seeding and domestic seeding events. Assuming international measles cases enter the county uniformly during their infectious period, the probability of onward transmission following a single imported case is

$$q_{0,c} = \int_0^1 q\left(0|k_M, \frac{k_M}{k_M + R_{E,c}x}\right)\mathrm{d}x \qquad [21]$$

where $q(x|k,p) = \frac{\Gamma(k+x)}{\Gamma(k)\Gamma(x+1)}p^k(1 - p)^x$ is the negative binomial distribution, $k_M$ is the dispersion coefficient for transmission from a single imported case, and $R_{E,c}$ is the effective reproduction number for county $c$.

Domestic seeding is assumed to occur proceeding outbreaks stemming from international imported cases. The expected number of cases in a county having internationally imported cases is

$$D_c = \left(1 - q_{0,c}^{I_c}\right)\frac{F_c}{N_c} \qquad [22]$$

where $I_c$ is the number of imported international measles cases into the county. We modeled the connectivity between counties using a gravity model of the form

$$Z_{c,j} = \log\left(N_c\right) + \log\left(N_j\right) - \lambda_\Delta \log\left(\delta_{c,j}\right) \qquad [23]$$

where $N_c$ is the population for county $c$, $\delta_{c,j}$ is the distance between the centroids of county $c$ and county $j$, and $\lambda_\Delta$ is the coefficient for distance.

The probability that county $c$ does not seed an outbreak in county $j$ is expressed by

$$\varrho_{c,j} = \exp\left\{-\lambda_\sigma D_c w_{c,j}\left(1 - q_{0,j}\right)\right\}, \qquad [24]$$

where $\lambda_\sigma$ is the domestic rate of seeding an outbreak and $w_{c,j} = \exp\left\{Z_{c,j}\right\}$. Thus, the probability that an outbreak is not seeded in county $j$ from other counties is expressed by

$$\tilde{\varrho}_j = \prod_c \varrho_{c,j}. \qquad [25]$$

Thus, the probability of zero annual local cases reported in county $c$ is expressed by

$$\tilde{q}_{0,c} = \varrho_c \tilde{q}_{0,c}^{I_c}. \qquad [26]$$

For $R_{E,c}$>1, the probability of observing $x$ measles cases in county $c$ for the year is expressed by

$$f\left(x|F_c, R_{S,s}\right) = \left(1 - \tilde{q}_{0,c}\right)h\left(x - 1|\tilde{N}_c, \Phi_c, r\right), \qquad [27]$$

where

$$h\left(x|\tilde{N}, \Phi, r\right) = \left(\frac{\Gamma(x+r)}{\Gamma(x+1)\Gamma(r)}\frac{\Gamma(N-r-x+1)}{\Gamma(\Phi-x+1)\Gamma(N-r-\Phi+1)}\right)/\left(\frac{\Gamma(N+1)}{\Gamma(\Phi+1)\Gamma(N-\Phi+1)}\right) \qquad [28]$$

is based on the formulation of a negative hypergeometric distribution, $\Phi_c = \left(\sum_a N_{c,a}\left(1 - \varphi_{c,a}\right)\right) - 1$ is the maximum outbreak size, $r$ is the hyperparameter for the distribution, and

$$\tilde{N}_c = \frac{\left[r\Phi_c + \left(F_c - 1\right)\phi_c - \left(F_c - 1\right)\right]}{\left(F_c - 1\right)} \qquad [29]$$

is the maximum number of elements. We note that we are using the distribution $h\left(x, \tilde{N}, \Phi, r\right)$ with support for $x \in \{0, \cdots, \Phi\}$ as the truncated distribution for the number of locally acquired cases. If $F_c < 1 + 10^{-8}$ and $R_{E,c}$>1 then we set $F_c = 1 + 10^{-8}$. This assumption allows for the computation of $\tilde{N}$. This approach is why we have subtracted one from the maximum outbreak size and the number of locally acquired cases. In the Monte Carlo sampling, we added one after the sample from this distribution to adjust for this assumption.

For $R_{E,c}$<1,the probability of observing $x$ measles cases in county $c$ for the year is expressed by

$$f\left(x|F_c, R_{S,s}\right) = \left(1 - \tilde{q}_{0,c}\right)s\left(x|R_E, k_M\right), \qquad [30]$$

where

$$s\left(x|R_E, k_M\right) = \frac{\Gamma\left(k_M x + x - 1\right)}{\Gamma\left(k_M x\right)\Gamma(x+1)}\left(\frac{R_E}{k_M}\right)^{x-1}\left(\frac{k_M}{k_M + R_E}\right)^{k_M x + x - 1} \qquad [31]$$

is the probability of a stuttering chain of size of at least one, i.e., $x \geq 1$ (9). We assumed that the size of a stuttering chain could not exceed 100 cases, i.e., if $x > 100$ then $s\left(x|R_E, k_M\right) = 0$. In addition, the source of the measles outbreak in Gaines county Texas is unknown (11). We assumed that three importation events occurred in this count, based on our own preliminary estimates of the effective reproductive number (attained through regression of the transmission rate) such that the likelihood of local transmission occurring was 0.50.

*Parameterization of vaccine coverage model*

We trained and validated our vaccination models for county- and state-level MMR vaccine coverage among kindergarteners spanning the 2017–18 school year through the 2023–24 school year (**File S1**). In addition, we trained the model using vaccine coverage data available at a health district level in Nebraska. Since we are only interested in inferring missing county-level MMR coverage data for the 2023–24 school year, validation was conducted only spatially, not temporally. We stratified the validation data into four groups based on the US Census Bureau's stratification: (1) Northeast (Connecticut, Maine, Massachusetts, New Hampshire, New Jersey, New York, Pennsylvania, Rhode Island, and Vermont), (2) Midwest (Illinois, Indiana, Iowa, Kansas, Michigan, Minnesota, Missouri, Nebraska, North Dakota, Ohio, South Dakota, and Wisconsin), (3) South (Alabama, Arkansas, Delaware, District of Columbia, Florida, Georgia, Kentucky, Louisiana, Maryland, Mississippi, North Carolina, Oklahoma, South Carolina, Tennessee, Texas, Virginia, and West Virginia) and (4) West (Arizona, California, Colorado, Idaho, Montana, Nevada, New Mexico, Oregon, Utah, Washington, and Wyoming). (12)

The coefficients of the regression model were estimated to maximize the log-likelihood

$$L_{MMR} = L_{County\,MMR} + L_{State\,MMR} + L_{NE\,MMR} + L_{Public} + L_{Uninsured} \quad [32]$$

where $L_{County\,MMR}$ is the log-likelihood for the county-level MMR coverage among kindergarteners, $L_{State\,MMR}$ is the log-likelihood for the state-level MMR coverage among kindergarteners, $L_{NE\,MMR}$ is the log-likelihood for the Nebraska health district MMR coverage among kindergarteners, $L_{Public}$ is the log-likelihood for the odds of vaccination of children that have public insurance relative to those with private insurance, and $L_{Uninsured}$ is the log-likelihood for the odds of vaccination of children that are uninsured relative to those with private insurance.

The log-likelihood for the county-level MMR coverage among kindergarteners is computed by

$$L_{County\,MMR} = \sum_{y=2017}^{2023} \sum_{c \in \vec{C}} \log\left(\eta\left(\log\left(\frac{\psi_{c,y}}{1-\psi_{c,y}}\right) \middle| \log\left(\frac{\nu_{c,y}}{1-\nu_{c,y}}\right), \sigma_{County}\right)\right) \quad [33]$$

where

$$\eta(x|\mu,\sigma) = \frac{1}{\sigma\sqrt{2\pi}}\exp\left\{-\frac{(x-\mu)^2}{2\sigma^2}\right\} \quad [34]$$

is the normal distribution with mean μ and standard deviation σ, $\psi_{c,y}$ is the reported vaccine coverage among kindergarteners for county $c$ in year $y$, $\nu_{c,y}$ is the model estimated vaccine coverage among kindergarteners for county $c$ in year $y$, $\vec{C}$ is the set of counties included in the training set, and $\sigma_{County}$ is the county-level hyper-parameter for the normal distribution.

The log-likelihood for the state-level MMR coverage among kindergarteners is computed by

$$L_{State\ MMR} = \sum_{y=2017}^{2023} \sum_{s \in \vec{S}} \log\left(\eta\left(\log\left(\frac{\Psi_{s,y}}{1-\Psi_{s,y}}\right) \mid \log\left(\frac{V_{s,y}}{1-V_{s,y}}\right), \sigma_{State}\right)\right) \quad [35]$$

where $\Psi_{s,y}$ is the reported vaccine coverage among kindergarteners for state $s$ in year $y$, $V_{s,y}$ is the model estimated vaccine coverage among kindergarteners for state $s$ in year $y$, $\vec{S}$ is the set of states included in the training set, and $\sigma_{State}$ is the state-level hyper-parameter for the normal distribution.

The log-likelihood for the Nebraska health district MMR coverage among kindergarteners is computed by

$$L_{NE\ MMR} = \sum_{y=2017}^{2023} \sum_{d \in \vec{D}} \log\left(\eta\left(\log\left(\frac{\overline{\Psi}_{d,y}}{1-\overline{\Psi}_{d,y}}\right) \mid \log\left(\frac{\overline{V}_{d,y}}{1-\overline{V}_{d,y}}\right), \sigma_{NE}\right)\right) \quad [36]$$

where $\overline{\Psi}_{d,y}$ is the reported vaccine coverage among kindergarteners for the Nebraska health district $d$ in year $y$, $\overline{V}_{d,y}$ is the reported vaccine coverage among kindergarteners for the Nebraska health district $d$ in year $y$, $\vec{D}$ is the set of districts included in the training set, and $\sigma_{NE}$ is the Nebraska health district level hyper-parameter for the normal distribution.

The log-likelihood for the odds of vaccination of children that have public insurance relative to those with private insurance

$$L_{Public} = \sum_{y=2017}^{2023} \sum_{s \in \vec{S}} \log\left(g\left(\theta_{s,y} \mid \alpha_{Public}, \frac{\Theta_{s,y}}{\alpha_{Public}}\right)\right) \quad [37]$$

where

$$g(x|a, b) \frac{1}{b^{a}\Gamma(a)} x^{a-1} \exp\left\{-\frac{x}{b}\right\} \quad [38]$$

is the Gamma distribution with the shape parameter $a$ and scale parameter $b$, $\theta_{s,y}$ is the reported odds of vaccination of a child publicly insured relative to a child who is privately insured in state $s$ for year $y$, and

$$\Theta_{s,y} = \left(\Psi^{M}_{s,y}\left(1 - \Psi^{P}_{s,y}\right)\right) / \left(\Psi^{P}_{s,y}\left(1 - \Psi^{M}_{s,y}\right)\right) \quad [39]$$

is the model estimated odds of vaccination of a child publicly insured relative to a child who is privately insured in state $s$ for year $y$, where $\Psi^{M}_{s,y}$ is the vaccine coverage among kindergarteners who are publicly insured in state $s$ in year $y$ and $\Psi^{P}_{s,y}$ is the vaccine coverage among kindergarteners who are privately insured in state $s$ in year $y$.

Similarly, the log-likelihood for the odds of vaccination of children that are uninsured relative to those with private insurance

$$L_{Uninsured} = \sum_{y=2017}^{2023} \sum_{s \in \vec{S}} \log\left(g\left(\tilde{\theta}_{s,y} | \alpha_{Uninsured}, \frac{\tilde{\Theta}_{s,y}}{\alpha_{Public}}\right)\right) \quad [40]$$

where $\alpha_{Uninsured}$ is the hyper-parameter for the Gamma distribution, $\tilde{\theta}_{s,y}$ is the reported odds of vaccination of a child uninsured relative to a child who is privately insured in state $s$ for year $y$, and

$$\tilde{\Theta}_{s,y} = \left(\Psi^{U}_{s,y}\left(1 - \Psi^{P}_{s,y}\right)\right) / \left(\Psi^{P}_{s,y}\left(1 - \Psi^{U}_{s,y}\right)\right) \quad [41]$$

is the model estimated odds of vaccination of a child publicly insured relative to a child who is privately insured in state $s$ for year $y$, where $\Psi^{M}_{s,y}$ is the vaccine coverage among kindergarteners who are publicly insured in state $s$ in year $y$ and $\Psi^{U}_{s,y}$ is the vaccine coverage among kindergarteners who are uninsured in state $s$ in year $y$.

*Parameterization of annual measles cases*

We used geographical and national measles cumulative incidence reported for 2025 (as of December 31, 2025) to train the model predicting the annual number of measles cases (13).

We estimated the distance coefficient $\lambda_{\Delta}$ for the gravity model, the dispersion coefficient $k_{M}$ for the transmission of measles from a single case, the domestic seeding rate $\lambda_{\sigma}$, the hyper-parameter $r$ for the county-level outbreak distribution, and the coefficients for the functional form of the transmission rate by maximizing the log-likelihood

$$L_{Outbreak} = L_{County} + L_{National} + L_{k}. \quad [42]$$

For cases reported within counties as of December 31, 2025 (14), the log-likelihood for county $c$ with $R_{E,c} > 1$ is expressed by

$$L_{c} = \log\left(\left(1 - \tilde{q}_{0,c}\right) h\left(x - 1 | \tilde{N}_{c}, \Phi_{c}, r\right)\right). \quad [43]$$

Similarly, the log-likelihood for county $c$ with $R_{E,c} < 1$ is expressed by

$$L_{c} = \log\left(\left(1 - \tilde{q}_{0,c}\right) s\left(x | R_{E,c}, k_{M}\right)\right). \quad [44]$$

For counties with no cases reported, the log-likelihood is expressed by

$$L_c = \log\left(\tilde{q}_{0,c}\right). \quad [45]$$

For cases with only regional information about the location (e.g., state-level or health districts consisting of multiple counties), the log-likelihood for county $c$ with $R_{E,c} > 1$ and at least one known case in the county (i.e., $x \geq 1$) is expressed by

$$L_c = \log\left(\sum_{u=0}^{U_{R,c}} b\left(u|U_{R,c}, \varsigma_c\right)\left(1 - \tilde{q}_{0,c}\right)h\left(x + u - 1|\tilde{N}_c, \Phi_c, r\right)\right), \quad [46]$$

where $U_{R,c}$ is the number of cases in the region containing county $c$ with no reported county location, $\varsigma_c$ is the susceptible population associated weight of the county relative to other counties in the region, and $b\left(u|U_{R,c}, \varsigma_c\right)$ is the Binomial distribution. Similarly, the log-likelihood for county $c$ with $R_{E,c} < 1$ and at least one known case in the county (i.e. $x \geq 1$) is expressed by

$$L_c = \log\left(\sum_{u=0}^{U_{R,c}} b\left(u|U_{R,c}, \varsigma_c\right)\left(1 - \tilde{q}_{0,c}\right)s\left(x + u|R_{E,c}, k_M\right)\right). \quad [47]$$

The log-likelihood for county $c$ with $R_{E,c} > 1$ and no known case in the county is expressed by

$$L_c = \log\left(b\left(0|U_{R,c}, \varsigma_c\right)\tilde{q}_{0,c} + \sum_{u=1}^{U_{R,c}} b\left(u|U_{R,c}, \varsigma_c\right)\left(1 - \tilde{q}_{0,c}\right)h\left(u - 1|\tilde{N}_c, \Phi_c, r\right)\right). \quad [48]$$

Similarly, the log-likelihood for county $c$ with $R_{E,c} < 1$ and no known case in the county is expressed by

$$L_c = \log\left(b\left(0|U_{R,c}, \varsigma_c\right)\tilde{q}_{0,c} + \sum_{u=1}^{U_{R,c}} b\left(u|U_{R,c}, \varsigma_c\right)\left(1 - \tilde{q}_{0,c}\right)s\left(u|R_{E,c}, k_M\right)\right). \quad [49]$$

If a county was included in an additional region, then the log-likelihood for county $c$ with $R_{E,c} > 1$ and at least one known case in the county (i.e., $x \geq 1$) is expressed by

$$L_c = \log\left(\sum_{u=0}^{U_{R,c}}\sum_{y=0}^{Y_{R,c}} b\left(u|U_{R,c}, \varsigma_c\right)b\left(y|Y_{R,c}, \hat{\varsigma}_c\right)\left(1 - \tilde{q}_{0,c}\right)h\left(x + u + y - 1|\tilde{N}_c, \Phi_c, r\right)\right), \quad [50]$$

and the log-likelihood for county $c$ with $R_{E,c} < 1$ and at least one known case in the county (i.e. $x \geq 1$) is expressed by

$$L_c = \log\left(\sum_{u=0}^{U_{R,c}} \sum_{y=0}^{Y_{R,c}} b\left(u|U_{R,c}, \varsigma_c\right) b\left(y|Y_{R,c}, \hat{\varsigma}_c\right)\left(1 - \tilde{q}_{0,c}\right) s\left(x + u + y|R_{E,c}, k_M\right)\right). \quad [51]$$

The log-likelihood for county $c$ with $R_{E,c} > 1$ and no known case in the county is expressed by

$$L_c = \log\{b\left(0|U_{R,c}, \varsigma_c\right) b\left(0|Y_{R,c}, \hat{\varsigma}_c\right)\tilde{q}_{0,c} + \sum_{u=1}^{U_{R,c}} \sum_{y=0}^{Y_{R,c}} b\left(u|U_{R,c}, \varsigma_c\right) b\left(y|Y_{R,c}, \hat{\varsigma}_c\right)\left(1 - \tilde{q}_{0,c}\right) h\left(u + y - 1|\tilde{N}_c, \Phi_c, r\right)$$

$$+ \sum_{u=0}^{U_{R,c}} \sum_{y=1}^{Y_{R,c}} b\left(u|U_{R,c}, \varsigma_c\right) b\left(y|Y_{R,c}, \hat{\varsigma}_c\right)\left(1 - \tilde{q}_{0,c}\right) h\left(u + y - 1|\tilde{N}_c, \Phi_c, r\right)\} \quad [52]$$

Similarly, the log-likelihood for county $c$ with $R_{E,c} < 1$ and no known case in the county is expressed by

$$L_c = \log\{b\left(0|U_{R,c}, \varsigma_c\right) b\left(0|Y_{R,c}, \hat{\varsigma}_c\right)\tilde{q}_{0,c} + \sum_{u=1}^{U_{R,c}} \sum_{y=0}^{Y_{R,c}} b\left(u|U_{R,c}, \varsigma_c\right) b\left(y|Y_{R,c}, \hat{\varsigma}_c\right)\left(1 - \tilde{q}_{0,c}\right) s\left(u + y|\tilde{N}_c, \Phi_c, r\right)$$

$$+ \sum_{u=0}^{U_{R,c}} \sum_{y=1}^{Y_{R,c}} b\left(u|U_{R,c}, \varsigma_c\right) b\left(y|Y_{R,c}, \hat{\varsigma}_c\right)\left(1 - \tilde{q}_{0,c}\right) s\left(u + y|\tilde{N}_c, \Phi_c, r\right)\} \quad (53)$$

Thus, the log-likelihood for the county-level incidence is expressed by

$$L_{County} = \frac{1}{A}\sum_c L_c, \quad [53]$$

where $A$ is the number of counties with three or more known cases, i.e., an outbreak in a county.

The log-likelihood for the national incidence is expressed by

$$L_{National} = \log\left(\kappa\left(\gamma_w|M\right)\right), \quad [54]$$

where κ is a kernel distribution for the sampled national incidence $M$ from the Monte Carlo simulations conducted during the optimization.

The dispersion parameter $k_M$ was restricted by the prior

$$L_k = \log\left(g\left(k_M | 11.53, 0.022\right)\right), \quad [55]$$

where the Gamma distribution has a maximum likelihood estimate of 0.23 with the shape value $11.53$ determined through least squares minimization of the 5th and 95th percentiles fit to the 90% CI of (0.16,0.39) (15).

We validated our modeling framework by reproducing national case counts reported in 2023 and 2024 using year-specific importation patterns (16), while assuming constant vaccination coverage (**Figure S2B and S2C; Table S2).**

*Reduction in MMR vaccine coverage*

We examined annual reductions in MMR vaccine coverage among children aged 0–6 years over 5 years. To approximate reductions in the age-specific groups 0–4, 5–9, and 10–14 years, we assumed an age-dependent exponential decline from the baseline vaccine coverage. For this approximation, we used single-age populations from 0 to 14 years. Single-age vaccine coverage was applied to 0–4 years, and coverage was uniform within the other age groups. For an annual reduction $\varepsilon$ in year $y$, we approximated the reductions in MMR vaccine coverage among single age group $a \in \{1, \cdots, 6 + (y - 1)\}$ by

$$\underline{\nu}_a = \nu_a / \left(1 + \exp\left\{-\chi_y((6 + y) - a)\right\}\right). \quad [56]$$

Thus, there is a reduction in vaccine coverage only among those 0–6 years of age in the first year and 0–11 years of age in the fifth year. All other age groups maintain their baseline vaccine coverage. Since a child is scheduled to receive their first dose at 12 months, those that are age 0, i.e., newborns, have a coverage level of zero, $\nu_0 = 0$. The value of $\chi_y$ is calibrated such that the reduction among those 0 to 6 years of age in year $y$ such that

$$\sum_{a=0}^{6} \nu_a \omega_a - \sum_{a=0}^{6} \underline{\nu}_a \omega_a = y\varepsilon, \quad [57]$$

where $\omega_a$ is the population associated weight for a child from those 0–6 years of age. Using these adjusted vaccine coverage levels and the population associated weights within the specified age groups, we computed the reduction within the entire age group.

Then within the specific age group, we computed the reduced county level coverage by adjusting the logistical transform of the county level coverage by $\Delta\mu$

$$\log\left(\tilde{\nu}_{U,c} / \left(1 - \tilde{\nu}_{U,c}\right)\right) = \beta_U + \sum_{j=1}^{N} \beta_j X_{j,c} + \Delta\mu \quad [58]$$

$$\log\left(\tilde{\nu}_{P,c} / \left(1 - \tilde{\nu}_{P,c}\right)\right) = \beta_P + \sum_{j=1}^{N} \beta_j X_{j,c} + \Delta\mu \quad [59]$$

$$\log\left(\tilde{\nu}_{M,c}/\left(1-\tilde{\nu}_{M,c}\right)\right)=\beta_M+\sum_{j=1}^{N}\beta_j X_{j,c}+\Delta\mu \quad [60]$$

such that we achieved the national level of reduction in the age group.

*Hospitalization, Severe Disease, and Death*

Using age- and vaccine status specified probabilities of hospitalization, severe disease, and death (**Table S10**), the number of hospitalizations, cases of severe disease, and number of measles-related deaths for a specified Monte Carlo sample was computed by the product of the probability and the case count. These values reflect the average number of hospitalizations, cases of severe disease, and number of measles-related deaths, which is more computationally feasible than running additional Monte Carlo samples for a single ensemble.

*Costs*

Costs associated with the outbreak response were broken down in cost associated with contact tracing, vaccinating contacts who are unvaccinated, and testing costs and presented in 2025 USD (**Table S1**). The number of contacts for a case in age group $a$ was computed by

$$\mathcal{C}_a=8\sum_{\forall j}M_{a,j} \quad [61]$$

where $\sum_{\forall j}M_{a,j}$ is the total number of daily contacts for an individual in age group $a$ and tracing is conducted for those exposed to the individual four days prior to four days after rash onset. Among the contacts, we assumed that 17.1% receive testing (17). The cost of testing is broken down into labour and materials (**Table S1**). The number of unvaccinated contacts for a case in age group $a$ was computed by

$$\mathcal{U}_a=8\sum_{\forall j}M_{a,j}\left(1-\nu_j\right), \quad [62]$$

where $\nu_j$ is the vaccine coverage among those in age group $j$. We assumed that only 13.8% of unvaccinated contacts receive the two doses of MMR required for post-exposure prophylaxis (18). Among those vaccinated under the age of 20, we assumed that 54% were eligible for the Vaccine For Children program (19).

Direct medical costs were associated with inpatient and outpatient costs. The probability of hospitalization was age and vaccine-status dependent (**Table S10**) (20). The duration of hospitalization was 3.84 days for cases 0 to 9 years of age, 5.52 days for cases 10 to 19 years of age, 5.33 days for cases 20 to 39 years of age and 5.44 days for cases 40 years of age or older (21). The average daily cost of a hospital stay varied by state, ranging from $1,088.78 in Mississippi to $4,717.00 in California, with the average daily cost of a hospital stay nationally being $3,304.32 (21, 22). With the absence of available state-level outpatient costs, we assumed a homogeneous cost of $408.08 per non-hospitalized case (17). We assumed these costs were the baseline costs for those privately insured. Commercial prices have been estimated to be roughly 2.5 times that for Medicare (23). For those 65 years of age and publicly insured, we applied state-level adjustment factors for inpatient and outpatient costs (23). Utilizing the fold-change hospital cost between Medicare and private insurance and the Medicaid-to-Medicare fee index (23, 24), we estimated the medical costs for those under the age of 65 and publicly insured. We assumed that those with private insurance are charged 10.7% more than those who are uninsured (25).

Productivity losses were computed based on lost wages from work and missed time at school. For cases under 15 years of age, we assumed a ratio of one caregiver to one case for time missed at work. Cases under the age of 20 were assumed to miss school and cases 15 years of age or older missed time at work. Cases under the age of 15 miss on average 7.3 days of school, while cases 15 years of age and older miss an average of 10.1 days of work or school (26). The economic value of missing a day of school was quantified by the daily education total public spending per pupil (27), with a daily value ranging from $25.72 in Idaho to $91.61 in New York. The daily average wage varied across counties (28).

For unvaccinated contacts under 15 years of age, we assumed a ratio of one caregiver to three unvaccinated contacts under 15 years of age for time missed at work (17). Unvaccinated contacts underwent a 21 day quarantine after exposure to a case (29).

*Monte Carlo Samples*

We conducted 2,500 Monte-Carlo samples. Prior to the optimization, we fixed the random number seed and sampled the maximum required size of uniform random numbers required. Utilizing the cumulative distribution functions and the uniform random numbers, we obtained the sampled outbreak size for the county. This approach allows for reproducibility of the likelihood for a specified parameter set.

Similarly to the optimization process, we fixed the random number seed and sampled the maximum required size of uniform random numbers required. Utilizing the cumulative distribution functions and the uniform random numbers, we obtained the sampled outbreak size for the county. This approach allows for reproducibility of a specified parameter set and a direct comparison of scenarios when the vaccine coverage is altered.

The final outbreak size among the counties where $R_{E,c} \leq 1$, (i.e. stuttering chain) was assumed to be no larger than 100 cases (10).

### Supplementary Figures

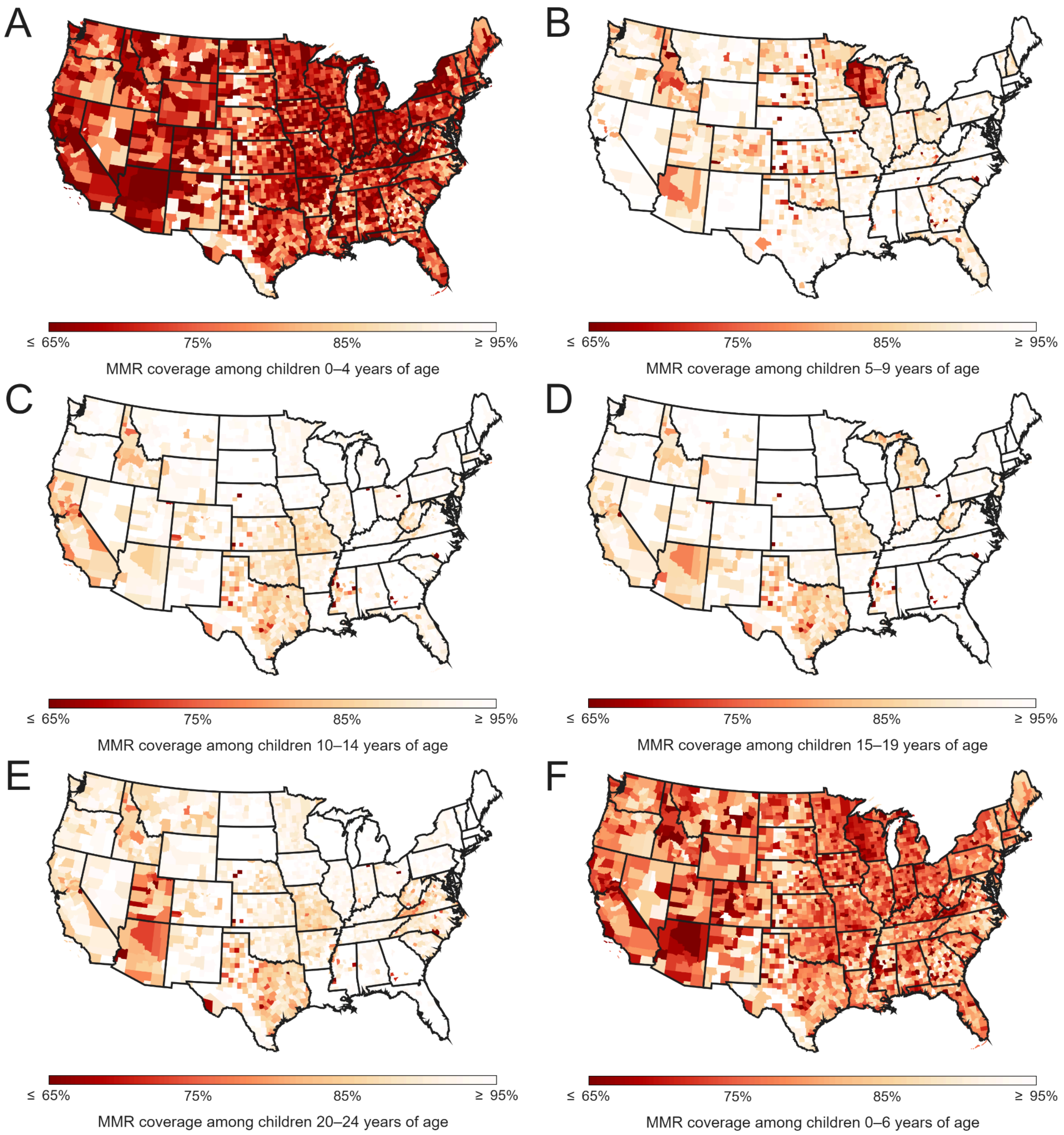


**Figure S1. County-level estimates of MMR coverage.** The estimated baseline county-level MMR coverage among (A) children 0–4 years of age, (B) children 5–9 years of age, (C) children 10–14 years of age, (D) children 15–19 years of age, (E) children 20–24 years of age, and (F) children 0–6 years of age based on 2023 vaccine coverage data. The coverage for children 0 years of age is specified to be 0%.

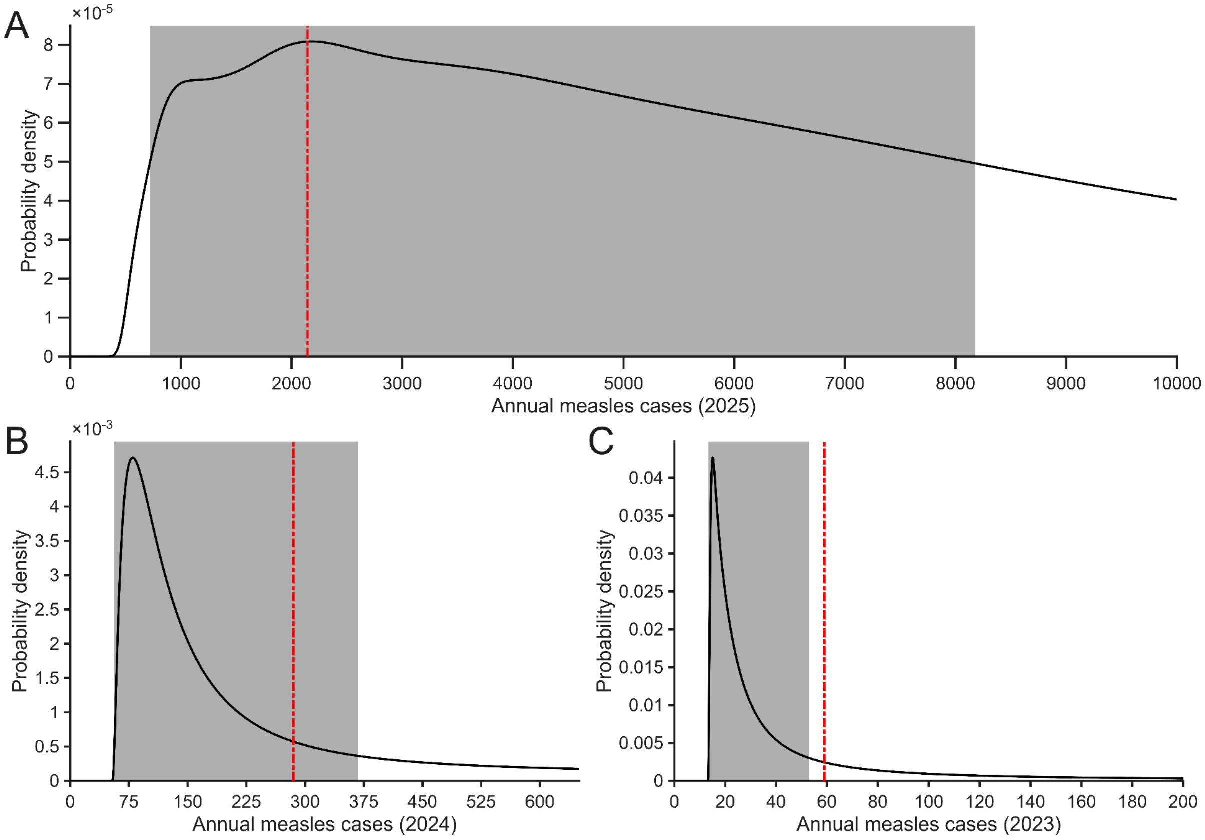


**Figure S2. Estimated annual measles incidence.** The estimated distribution (solid black line) and 50% high density interval (gray area) of annual measles incidence for the United States compared to the reported measles cases (red-dashed line) in (A) 2025, (B) 2024, and (C) 2023 based on the number of importation events reported for the year.

### Supplementary Tables

**Table S1.** Summary of costs (2025 USD) applied to outbreak response, direct medical, and productivity losses

| Category | Measure | | Value | Reference |
|---|---|---|---|---|
| Outbreak response | Contact tracing per contact | | $559.45 | (17) |
| | Two doses of MMR to unvaccinated contacts who have private insurance or are over 19 years of age | | $190.40 | (30) |
| | Two doses of MMR to unvaccinated contacts who do not have private insurance and are under 20 years of age | | $52.66 | (30) |
| | Administration fee for two doses of MMR to unvaccinated contacts who have private insurance or are over 19 years of age | | $51.60 | (31) |
| | Administration fee for two doses of MMR to unvaccinated contacts who do not have private insurance and are under 20 years of age | | $26.50 | (31) |
| | Labour cost per test | | $191.92 | (17) |
| | Material cost per test | | $29.45 | (17) |
| Direct medical | National average hospitalization cost(Varies by state) | Under 10 years of age | $12,688.59 | (21, 22) |
| | | 10–19 years of age | $18,239.85 | (21, 22) |
| | | 20–39 years of age | $17,612.03 | (21, 22) |
| | | 40 years of age or older | $17,975.50 | (21, 22) |
| | Cost for a non-hospitalized case | | $408.08 | (17) |
| Productivity losses | National average associated productivity loss for a case of age (varies by county) | under 15 years | $1,553.88 | (26, 32) |
| | | 15 years of age or older | $2,149.89 | (26, 32) |
| | National average associated productivity loss for a case attributed to school absenteeism (varies by state) | Under 15 years of age | $345.54 | (26, 27) |
| | | 15–19 years | $478.08 | (26, 27) |
| | National average associated productivity loss per contact (varies by county) | Under 15 years of age | $1,490.02 | (17, 29, 33) |
| | | 15 years of age or older | $4,470.06 | (29, 34) |
| | National average associated productivity loss for a contact under 20 years of age attributed to school absenteeism (varies by state) | | $994.02 | (27, 29) |

**Table S2.** The mode and 50% high-density interval of the estimated annual measles cases, hospitalizations, costs, and costs per case for 2023 to 2025 from 2,500 Monte Carlo simulations.[a]

| Measure | Estimated value | | |
|---|---|---|---|
| | 2025 | 2024 | 2023 |
| Cases | 2,181 (719–8,176) | 80 (56–368) | 15 (13–53) |
| Hospitalizations | 554 (196–1,927) | 25 (9–99) | 5 (2–11) |
| Severe disease | 390 (138–1,375) | 18 (7–70) | 3 (2–8) |
| Deaths | 5 (2–17) | 0 (0–1) | 0 (0–0) |
| Costs | $244.2 million ($69.9–$872.5 million) | $8.5 million ($2.9–$37.1 million) | $1.6 million ($0.8–$4.0 million) |
| Outbreak response | $144.3 million ($44.2–$556.5 million) | $5.1 million ($1.7–$23.6 million) | $0.97 million ($0.52–$2.39 million) |
| Direct medical | $5.9 million ($2.5–$24.4 million) | $0.35 million ($0.14–$1.26 million) | $0.07 million ($0.03–$0.16 million) |
| Uninsured | $1.87 million ($0.60–$5.69 million) | $0.06 million ($0.02–$0.26 million) | $0.01 million ($0.00–$0.04 million) |
| Public | $0.68 million ($0.35–$3.27 million) | $0.05 million ($0.02–$0.19 million) | $0.01 million ($0.00–$0.02 million) |
| Private | $3.01 million ($1.43–$14.92 million) | $0.24 million ($0.09–$0.80 million) | $0.04 million ($0.02–$0.11 million) |
| Productivity losses | $84.4 million ($23.0–$286.2 million) | $3.0 million ($1.1–$12.1 million) | $0.55 million ($0.29–$1.40 million) |
| Cost per case | $104,629 ($100,729–$110,140) | $97,336 ($93,895–$102,310) | $97,740 ($94,015–$105,760) |

[a] The modes of the cost may not sum up to one, as the outcomes reflect joint distributions of outbreak response cost, direct medical, and productivity losses. Similarly, the mode of the cost per case may not reflect the mode of overall cost divided by the mode of the estimated number of cases

**Table S3.** The mode and 50% high-density interval of the estimated costs breakdown for 2023 to 2025 from 2,500 Monte Carlo simulations. [a]

| Measure | Estimated proportion of cost | | |
|---|---|---|---|
| | 2025 | 2024 | 2023 |
| Outbreak response | 65.21% (62.50%–66.92%) | 61.88% (60.05%–64.68%) | 61.86% (59.39%–64.63%) |
| Productivity loses | 32.07% (30.11%–34.80%) | 33.97% (31.93%–36.17%) | 34.21% (31.68%–36.58%) |
| Direct medical | 2.95% (2.66%–3.16%) | 3.82% (3.14%–4.16%) | 3.98% (3.24%–4.45%) |
| Uninsured [b] | 20.85% (18.66%–24.84%) | 19.62% (15.67%–23.72%) | 20.03% (14.31%–26.48%) |
| Public [b] | 14.22% (10.73%–15.37%) | 13.81% (11.42%–16.51%) | 10.59% (3.43%–13.86%) |
| Private [b] | 63.26% (57.73%–67.25%) | 65.02% (60.63%–70.84%) | 64.44% (57.52%–72.23%) |

[a] The modes may not sum up to one, as the outcomes reflect joint distributions of the proportion of the overall costs, and proportion of the total direct medical costs.

**Table S4.** The mode and 50% high-density interval of the annual measles cases in the US based on imported measles cases reported for 2025 with an absolute annual reduction in vaccine coverage among children from 0 to 6 years of age ranging from 0.25% per year to 1% per year.

| Year | Annual reduction in vaccine coverage among children from 0 to 6 years | | | |
|---|---|---|---|---|
| | 0.25% | 0.5% | 0.75% | 1% |
| 2025 | 2,181 (719–8,176) | 2,181 (719–8,176) | 2,181 (7,19–8,176) | 2,181 (719–8,176) |
| 2026 | 2,263 (755–8,367) | 2,299 (777–8,547) | 2,361 (8,21–8,763) | 4,057 (872–9,000) |
| 2027 | 2,313 (789–8,568) | 4,106 (881–9,054) | 4,426 (9,89–9,633) | 4,686 (1,126–10,330) |
| 2028 | 2,446 (863–8,894) | 4,469 (1,064–9,975) | 5,254 (1,558–11,617) | 5,891 (2,590–13,764) |
| 2029 | 4,261 (963–9,435) | 5,248 (1,638–11,778) | 6,293 (3,172–14,975) | 9,945 (5,256–18,774) |
| 2030 | 4,580 (1,135–10,237) | 5,914 (764–13,102) | 10,545 (5,693–19,753) | 17,232 (9,177–26,428) |

**Table S5.** The mode and 50% high-density interval of annual measles hospitalizations in the US based on imported measles cases reported for 2025 with an absolute annual reduction in vaccine coverage among children from 0 to 6 years of age ranging from 0.25% per year to 1% per year.

| Year | Annual reduction in vaccine coverage among children from 0 to 6 years | | | |
|---|---|---|---|---|
| | 0.25% | 0.5% | 0.75% | 1% |
| 2025 | 554 (196–1,927) | 554 (196–1,927) | 554 (196–1,927) | 554 (196–1,927) |
| 2026 | 582 (204–1,973) | 862 (211–2,017) | 937 (221–2,068) | 968 (232–2,124) |
| 2027 | 863 (213–2,021) | 982 (235–2,136) | 1,072 (264–2,278) | 1,122 (301–2,447) |
| 2028 | 942 (230–2,097) | 1,083 (283–2,356) | 1,253 (366–2,708) | 1,405 (627–3,243) |
| 2029 | 1,025 (255–2,225) | 1,252 (377–2,735) | 1,510 (770–3,531) | 2,363 (1,245–4,406) |
| 2030 | 1,098 (300–2,413) | 1,432 (726–3,420) | 2,504 (1,347–4,625) | 4,085 (2,184–6,210) |

**Table S6.** The mode and 50% high-density interval of annual measles deaths in the US based on imported measles cases reported for 2025 with an absolute annual reduction in vaccine coverage among children from 0 to 6 years of age ranging from 0.25% per year to 1% per year.

| Year | Annual reduction in vaccine coverage among children from 0 to 6 years | | | |
|---|---|---|---|---|
| | 0.25% | 0.5% | 0.75% | 1% |
| 2025 | 5 (2–17) | 5 (2–17) | 5 (2–17) | 5 (2–17) |
| 2026 | 5 (2–17) | 8 (2–18) | 8 (2–18) | 9 (2–18) |
| 2027 | 8 (2–18) | 9 (2–19) | 9 (2–20) | 10 (3–21) |
| 2028 | 8 (2–18) | 9 (2–20) | 11 (3–24) | 12 (6–28) |
| 2029 | 9 (2–19) | 11 (3–24) | 13 (7–31) | 21 (11–38) |
| 2030 | 10 (3–21) | 12 (6–30) | 22 (12–40) | 36 (19–54) |

**Table S7.** The mode and 50% high-density interval of the cost per measles case in the US based on imported measles cases reported for 2025 with an absolute annual reduction in vaccine coverage among children from 0 to 6 years of age ranging from 0.25% per year to 1% per year.

| Year | Annual reduction in vaccine coverage among children from 0 to 6 years | | | |
|---|---|---|---|---|
| | 0.25% | 0.5% | 0.75% | 1% |
| 2025 | $104,629 ($100,729–$110,140) | $104,629 ($100,729–$110,140) | $104,629 ($100,729–$110,140) | $104,629 ($100,729–$110,140) |
| 2026 | $104,729 ($100,715–$110145) | $104,778 ($100,679–$110,161) | $104,708 ($100,639–$110,079) | $104,695 ($100,598–$110,050) |
| 2027 | $104,663 ($100,693–$110176) | $104,565 ($100,575–$110,038) | $104,639 ($100,637–$110,091) | $104,460 ($100,624–$110,059) |
| 2028 | $104,754 ($100,697–$110145) | $104,531 ($100,760–$110,185) | $104,651 ($100,646–$110,160) | $104,816 ($100,948–$110,415) |
| 2029 | $104,771 ($100,763–$110196) | $104,600 ($100,828–$110,352) | $105,221 ($101,381–$110,750) | $106,531 ($101,838–$110,936) |
| 2030 | $104,621 ($100,908–$110333) | $105,190 ($101,389–$110,801) | $106,782 ($102,125–$111,184) | $107,723 ($102,999–$111,924) |

**Table S8.** The mode and 50% high-density interval of the cumulative costs (billions) attributed to measles cases in the US based on imported measles cases reported for 2025 with an absolute annual reduction in vaccine coverage among children from 0 to 6 years of age ranging from 0.25% per year to 1% per year.

| Year | Annual reduction in vaccine coverage among children from 0 to 6 years | | | |
|---|---|---|---|---|
| | 0.25% | 0.5% | 0.75% | 1% |
| 2026 | $0.25 ($0.07–$0.89) | $0.26 ($0.08–$0.91) | $0.27 ($0.08–$0.93) | $0.28 ($0.09–$0.96) |
| 2027 | $1.34 ($0.55–$2.41) | $1.42 ($0.60–$2.50) | $1.53 ($0.64–$2.59) | $1.60 ($0.68–$2.68) |
| 2028 | $2.40 ($1.24–$4.00) | $2.58 ($1.38–$4.25) | $2.70 ($1.57–$4.55) | $2.95 ($1.72–$4.86) |
| 2029 | $3.79 ($2.15–$5.81) | $4.19 ($2.44–$6.32) | $4.63 ($2.76–$6.93) | $5.10 ($3.14–$7.56) |
| 2030 | $4.92 ($3.18–$7.72) | $5.56 ($3.79–$8.75) | $6.63 ($4.49–$10.03) | $7.77 ($5.56–$11.58) |

**Table S9.** Summary of the covariates used in the logistic regression model for predicting the vaccine coverage among kindergarteners for the 2023–2024 school year

| Measure | Percent of counties | Reference |
|---|---|---|
| Philosophical exemptions | 35.2% | **File S1** |
| Religious exemptions | 93.1% | **File S1** |
| Rural Urban Continuum Code = 1 | 15.0% | (3) |
| Rural Urban Continuum Code = 2 | 12.4% | (3) |
| Rural Urban Continuum Code = 3 | 11.5% | (3) |
| Rural Urban Continuum Code = 4 | 6.3% | (3) |
| Rural Urban Continuum Code = 5 | 2.4% | (3) |
| Rural Urban Continuum Code = 6 | 12.0% | (3) |
| Rural Urban Continuum Code = 7 | 7.7% | (3) |
| Rural Urban Continuum Code = 8 | 14.5% | (3) |
| Rural Urban Continuum Code = 9 | 18.3% | (3) |

| Measure | Percentiles | | | Reference |
|---|---|---|---|---|
| | $50^{th}$ | $2.5^{th}$ | $97.5^{th}$ | |
| Physicians per 100,000 population | 4.50 | 0 | 13.58 | (1) |
| Percent of population: 20–24 | 5.59% | 2.80% | 12.24% | (2) |
| Percent of population: 25–29 | 5.76% | 3.36% | 8.57% | (2) |
| Percent of population: 30–34 | 5.92% | 3.55% | 8.54% | (2) |
| Percent of population: 35–39 | 6.04% | 3.82% | 8.51% | (2) |
| Percent of population: 40–44 | 5.97% | 3.73% | 8.27% | (2) |
| Percent of population: White | 83.27% | 26.43% | 96.05% | (2) |
| Percent of population: African American | 2.27% | 0.00% | 51.92% | (2) |
| Percent of population: American Indian and Alaskan Native | 0.31% | 0.00% | 14.02% | (2) |

| | | | | |
|---|---|---|---|---|
| Percent of population: Asian | 0.61% | 0.00% | 7.77% | (2) |
| Percent of population: Native Hawaiian and Pacific Islander | 0.01% | 0.00% | 0.69% | (2) |
| Median family income ($\log_{10}$) | 4.90 | 4.59 | 5.12 | (2) |
| Gini index | 0.445 | 0.384 | 0.531 | (2) |
| Percent of population: Less than grade 9 | 3.42% | 0.90% | 14.67% | (2) |
| Percent of population: Grade 9–12 | 6.57% | 2.29% | 14.35% | (2) |
| Percent of population: High school grad | 33.82% | 17.49% | 47.72% | (2) |
| Percent of population: Some College | 20.90% | 12.31% | 28.96% | (2) |
| Percent of population: Associate Degree | 9.55% | 4.73% | 16.40% | (2) |
| Percent of population: Bachelor Degree | 14.60% | 6.73% | 29.83% | (2) |
| Percent of population: Graduate/Professional Degree | 7.07% | 3.03% | 21.84% | (2) |
| Percent of population with poverty index ratio under 0.50 | 0.97% | 0.20% | 3.72% | (2) |
| Percent of population with poverty index ratio 0.50–0.74 | 0.58% | 0.04% | 2.20% | (2) |
| Percent of population with poverty index ratio 0.75–0.99 | 0.72% | 0.14% | 2.39% | (2) |
| Percent of population with poverty index ratio 1.00–1.24 | 0.87% | 0.19% | 2.36% | (2) |
| Percent of population with poverty index ratio 1.25–1.49 | 0.98% | 0.27% | 2.44% | (2) |
| Percent of population with poverty index ratio 1.50–1.74 | 1.05% | 0.31% | 2.36% | (2) |
| Percent of population with poverty index ratio 1.75–1.84 | 0.40% | 0.00% | 1.14% | (2) |
| Percent of population with poverty index ratio 1.85–1.99 | 0.64% | 0.13% | 1.56% | (2) |
| Percent of population with poverty index ratio 2.00–2.99 | 4.43% | 2.20% | 6.49% | (2) |

| | | | | |
|---|---|---|---|---|
| Percent of population with poverty index ratio 3.00–3.99 | 3.88% | 1.67% | 5.85% | (2) |
| Percent of population with poverty index ratio 4.00–4.99 | 3.07% | 0.95% | 4.77% | (2) |
| Percent of population with poverty index ratio 5.00 and over | 6.83% | 1.71% | 14.35% | (2) |

**Table S10.** Probability of hospitalization, severe disease, and death stratified by age and vaccine status

| Measure | Age group | Unvaccinated | Vaccinated | Reference |
|---|---|---|---|---|
| Hospitalization | 0 to 4 years of age | 0.283 | 0.204 | (35) |
| | 5 to 19 years of age | 0.116 | 0.111 | (35) |
| | 20 years of age or older | 0.397 | 0.191 | (35) |
| Severe disease | 0 to 4 years of age | 0.237 | 0.179 | (35) |
| | 5 to 19 years of age | 0.085 | 0.096 | (35) |
| | 20 years of age or older | 0.249 | 0.161 | (35) |
| Deaths | 0 to 4 years of age | 0.0025 | | (36) |
| | 5 to 19 years of age | 0.0011 | | (36) |
| | 20 years of age or older | 0.0029 | | (36) |

**Table S11.** The assumed priors and maximum likelihood estimate for the parameters in the measles incidence model

| | Parameter | Estimate | Prior |
|---|---|---|---|
| $\log_{10}(\lambda_\Delta)$ | Gravity model: Distance | -1.151 | $(-5, 1)$ |
| $\log_{10}(\lambda_\sigma)$ | Seeding rate | -8.946 | $(-16, -6)$ |
| $\Upsilon_{R,1}$ | Transmission rate: RUCC=1 | -11.412 | $(-27.873,\ 5.320)$[a] |
| $\Upsilon_{R,2}$ | Transmission rate: RUCC=2 | -11.161 | $(-27.816,\ 5.350)$[a] |
| $\Upsilon_{R,3}$ | Transmission rate: RUCC=3 | -10.734 | $(-27.705,\ 5.377)$[a] |
| $\Upsilon_{R,4}$ | Transmission rate: RUCC=4 | -11.286 | $(-27.687,\ 5.486)$[a] |
| $\Upsilon_{R,5}$ | Transmission rate: RUCC=5 | -11.225 | $(-27.507,\ 5.631)$[a] |
| $\Upsilon_{R,6}$ | Transmission rate: RUCC=6 | -10.962 | $(-27.800,\ 5.204)$[a] |
| $\Upsilon_{R,7}$ | Transmission rate: RUCC=7 | -11.028 | $(-27.793,\ 5.528)$[a] |
| $\Upsilon_{R,8}$ | Transmission rate: RUCC=8 | -11.633 | $(-27.775,\ 5.657)$[a] |
| $\Upsilon_{R,9}$ | Transmission rate: RUCC=9 | -10.903 | $(-27.710,\ 5.182)$[a] |
| $\Upsilon_1$ | Transmission rate:Median family income ($\log_{10}$) | 2.293 | $(-0.635,\ 5.384)$[a] |
| $\Upsilon_2$ | Transmission rate:Gini index | -3.886 | $(-6.465,\ -1.132)$[a] |
| $\Upsilon_3$ | Transmission rate:Percent of population: Grade 9–12 | 1.349 | $(-4.083,\ 6.667)$[a] |
| $\Upsilon_4$ | Transmission rate:Percent of population: High school grad | 1.690 | $(-0.338,\ 3.647)$[a] |
| $\Upsilon_5$ | Transmission rate:Percent of population: Some College | -1.179 | $(-3.252,\ 1.036)$[a] |
| $\Upsilon_6$ | Transmission rate:Percent of population: Associate Degree | 4.912 | $(0.941,\ 8.778)$[a] |
| $\Upsilon_7$ | Transmission rate:Percent of | 1.550 | $(-0.746,\ 3.909)$[a] |

| | | | |
|---|---|---|---|
| | population: Bachelor Degree | | |
| $\Upsilon_8$ | Transmission rate:Percent of population: Graduate /Professional Degree | 0.950 | $(-0.953, 5.194)$[a] |
| $\Upsilon_9$ | Transmission rate: Percent of population with poverty index ratio under 0.50 | 10.404 | $(-14.095, 32.618)$[a] |
| $\Upsilon_{10}$ | Transmission rate: Percent of population with poverty index ratio 0.50–0.74 | -2.406 | $(-33.989, 32.256)$[a] |
| $\Upsilon_{11}$ | Transmission rate: Percent of population with poverty index ratio 0.75–0.99 | 56.512 | $(34.094, 78.204)$[a] |
| $\Upsilon_{12}$ | Transmission rate: Percent of population with poverty index ratio 1.00–1.24 | 9.020 | $(-26.680, 46.161)$[a] |
| $\Upsilon_{13}$ | Transmission rate: Percent of population with poverty index ratio 1.25–1.49 | -15.338 | $(-42.793, 10.622)$[a] |
| $\Upsilon_{14}$ | Transmission rate: Percent of population with poverty index ratio 1.50–1.74 | -1.643 | $(-24.107, 16.696)$[a] |
| $\Upsilon_{15}$ | Transmission rate: Percent of population with poverty index ratio 1.75–1.84 | 26.118 | $(-13.388, 54.400)$[a] |
| $\Upsilon_{16}$ | Transmission rate: Percent of population with poverty index ratio 1.85–1.99 | 8.934 | $(-9.125, 27.680)$[a] |
| $\Upsilon_{17}$ | Transmission rate: Percent of population with poverty index ratio 2.00–2.99 | -0.633 | $(-8.893, 7.810)$[a] |
| $\Upsilon_{18}$ | Transmission rate: Percent of population with poverty index ratio 3.00–3.99 | -20.524 | $(-29.859, -10.772)$[a] |
| $\Upsilon_{19}$ | Transmission rate: Percent of population with poverty index ratio 4.00–4.99 | 7.796 | $(-1.927, 17.341)$[a] |

| | | | |
|---|---|---|---|
| $\Upsilon_{20}$ | Transmission rate: Percent of population with poverty index ratio 5.00 and over | -8.218 | $(-14.390, \ -2.174)$[a] |
| $k_M$ | Dispersion coefficient for negative binomial | 0.231 | Gamma distribution<br>$a = 11.53$<br>$b = 0.022$ (Ref (15)) |
| $r$ | Hyper-parameter | 1 | $\{1, \cdots, 30\}$ |

[a] The priors were formed from the 99% confidence intervals attained from an independent linear regression of only the county-level transmission rate among counties with at least three cases. In this regression analysis, we only used cases where the location was known.
